# Real-World Cuff-Measured Blood Pressure Trajectories in 20,003 Consumer Wearable Users

**DOI:** 10.64898/2026.11.17.26368311

**Authors:** C.P. Cheung, J.G. Cohen, A.B. Bhatt, D Curran, L.R. Parola, F. Fielding, C.J. Chapman, W. von Hippel, K.E. Holmes, G.J. Grosicki

## Abstract

Consumer wearables increasingly allow users to log cuff-measured blood pressure (BP), but resulting BP trajectories are uncharacterized. In this retrospective cohort study, 20,003 adults with Elevated BP or Stage 1 or Stage 2 Hypertension logged BP via a consumer wearable platform. We modeled 6-month trajectories with linear mixed-effects models and examined associations of ≥5 mmHg reduction with engagement and wearable-derived metrics. Adjusted systolic reductions ranged from −2.6 mmHg (95% CI −3.1 to −2.2) in Elevated BP to −12.1 mmHg (−12.5 to −11.7) in Stage 2 Hypertension; diastolic reductions were −3.7 and −9.2 mmHg in Stage 1 and Stage 2. Overall, 50.2% achieved a ≥5 mmHg systolic reduction and 27.9% reached normal BP. App time, journaling, AI Coach use, and BP logging were associated with higher odds of reduction; longer sleep and greater physical activity accompanied lower BP. These associational findings support consumer wearables as potentially scalable BP-monitoring platforms.

## Introduction

As the leading cause of mortality worldwide,^1^ cardiovascular disease (CVD) imposes a substantial public health burden. Hypertension is a major contributor to CVD, accounting for approximately half the risk of coronary heart disease and stroke^2^ and affecting more than 60% of U.S. adults aged 45–75.^3^ Although effective pharmacologic and lifestyle interventions for reducing blood pressure (BP) are well established,^4–6^ fewer than half of U.S. adults with hypertension achieve BP control.^7^ This disconnect highlights the need for approaches capable of supporting BP management at scale. Home BP monitoring is recommended by current guidelines for the diagnosis and management of hypertension,^8,9^ and home-measured BP predicts cardiovascular events independently of, and at least as strongly as, office readings.^10,11^

Remotely delivered interventions offer a potential solution for scalable BP control. Systematic reviews of randomized trials indicate that self-monitoring and telemonitoring of BP lower BP relative to usual care,^12^ although this benefit depends on coupling measurement with co-interventions such as feedback, counseling, or medication titration.^13^ Single-arm observational cohort studies of digital hypertension self-management programs combining BP monitoring with smartphone applications, lifestyle education, or coaching, have reported systolic BP (SBP) reductions of approximately 7–20 mmHg over 6 months to three years, with larger reductions among those with uncontrolled hypertension.^14–17^ Although a randomized trial of telephone-based health coaching and home BP monitoring in young adults with uncontrolled hypertension did not improve BP relative to usual care, it did improve behaviors relevant to BP control, including increased physical activity and reductions in dietary sodium.^18^ Importantly, greater engagement with digital BP monitoring programs has also been associated with greater BP reductions,^16^ suggesting that greater engagement may facilitate these benefits. Collectively, these findings provide evidence that remotely delivered interventions are associated with reduced BP and promotion of behaviors that facilitate long-term BP control.

While these remote interventions show promise, their reach is constrained by eligibility, health-plan coverage, healthcare access, and the demands of structured clinical or coaching infrastructure. Consumer wearables may offer a more accessible approach. Approximately two-in-five U.S. adults now use wearable devices, with high adoption among individuals at elevated cardiovascular risk.^19^ Similar to digital BP management programs, greater engagement with wearable platforms has been associated with favorable changes in behaviors and physiological markers relevant to BP control, including physical activity, sleep, resting heart rate, and heart rate variability.^20–22^ Wearable platforms have also begun integrating BP monitoring features, allowing users to track cuff-measured BP and creating the potential to examine BP trajectories alongside passive behavioral, physiological, and engagement data in real-world settings. However, whether engagement with a consumer wearable platform is directly associated with BP change remains unclear.

Accordingly, we sought to characterize BP trajectories among people using a consumer wearable platform, and identify behavioral, physiological, and digital engagement factors associated with BP change. We hypothesized that initiating BP tracking would be associated with reductions in BP, that wearable-derived behavioral and physiological data would associate with those changes, and that greater engagement with the digital platform would be associated with larger BP reductions.

## Methods

### Ethics Statement

This study was determined exempt from IRB review by Salus IRB (#26316) under 45 CFR 46.104(d)(4)(ii), as it involved secondary analysis of deidentified data.

### Study Design

We conducted a retrospective, real-world observational cohort study of adult users (age 18–79) of a consumer wearable device (WHOOP, Inc., Boston, USA). Devices were obtained through standard consumer channels (self-purchased or employer/organization-provided) and were not provided for the study. Participants were not randomized to WHOOP use or to any BP intervention. The WHOOP wearable was not used itself to measure BP. Instead, the platform provides an optional blood pressure tracking feature, where users self-report BP readings that they obtained themselves, using a separate cuff-based BP device (e.g., a home or clinic oscillometric or auscultatory cuff). All BP values analyzed here were cuff-measured on such external devices and manually entered by the user, and no photoplethysmography-derived or otherwise device-estimated BP values were used.

All BP measures required users to perform triplicate recordings, and thus, each BP measurement represents the average of three consecutive cuff measurements. At each triplicate manual cuff reading, users were provided guideline-recommended practices, specifically, to ensure at least five minutes of seated rest, no caffeine, food, smoking or exercise within two hours, arm supported at heart level, and feet flat with back supported, before readings. Cuff make, model and measurement setting were not available. Self-reporting of BP was voluntary. After each entry, users could see the averaged reading alongside previous readings. The platform otherwise provided its standard behavioral feedback, including a nightly sleep need, a daily physical activity target, notifications and an on-demand AI Coach, which encouraged adequate sleep and regular physical activity but was not tailored to BP. No BP-specific coaching, alerts, medication reminders or clinical follow-up were provided.

Users were eligible if they manually entered a cuff-measured reading on the platform and a subsequent cuff-measured reading 150–240 days later, with both readings occurring between April 19, 2025 and May 13, 2026. The first reading was defined as the user’s first-ever cuff-measured entry and served as the baseline, anchoring each participant’s study period. When multiple readings fell within the 150–240-day follow-up window, the one closest to 180 days was selected as the 6-month BP. To exclude implausible entries, values were retained only within 70–250 mmHg (systolic) and 40–150 mmHg (diastolic).

For further analyses, all individual cuff-measured BP measurements recorded in the 12 months after baseline were retained. The 12-month value was defined as the measurement closest to 365 days within a 335–390-day window. A 3-month value was defined as the measurement closest to 90 days within a 60–120-day window.

### Wearable Device & Digital Health Platform Derived Metrics

Wearable-derived sleep, cardio-autonomic, and physical activity metrics were calculated using previously described accelerometry- and photoplethysmography-based methods via the wearable device.^23–25^ Sleep duration was defined as the total time in bed spent asleep. Sleep consistency was quantified using a 0–100 score adapted from the Sleep Regularity Index.^26^ It evaluates sleep onset and offset times over a rolling window, comparing the most recent 24 hours to the preceding four days, with comparisons of days further apart assigned progressively lower weights,^27^ and higher scores indicating lower variability in sleep-wake timing. Physical activity was summarized using Strain, a logarithmic score (0–21) quantifying cardiovascular load using heart-rate intensity zones. Wearable-derived cardio-autonomic metrics included resting heart rate (RHR) and heart rate variability (HRV). Both were derived from non-wake periods of the primary sleep episode. RHR was calculated from beat-to-beat intervals, and HRV quantified as the root mean square of successive differences between heartbeats (RMSSD). WHOOP has been validated against polysomnography and electrocardiography.^28–30^

Application engagement and attainment of wearable-derived behavioral recommendations were quantified during the study period using average daily time in the application, proportion of study days with an entry in the platform’s daily behaviors journal, push notification engagement, artificial intelligence (AI) Coach interaction, proportion of study days recommended daily Strain target was met, average nightly sleep need achieved, and total number of triplicate BP entries recorded.

Health behaviors were captured through the in-app journal, including antihypertensive medication, GLP-1 receptor agonist, and tobacco or nicotine use. Antihypertensive medication use was defined by affirmative entries on either the "Blood Pressure Medication" or "Beta Blockers" tracker from 180 days before baseline through the 6-month reading. Participants were classified as established users (first log ≥ 8 weeks before baseline), recent initiators (within the 8 weeks before baseline), mid-study initiators (on or after baseline), or never-logging. For covariate adjustment, established users and recent initiators were classified as using antihypertensive medication at baseline. Owing to a smaller number of GLP-1 receptor agonist users, GLP-1 receptor agonist use was not included in model parameters and was instead used in sensitivity analyses.

### Statistical Analysis

The primary cohort included participants with baseline BP classified as Elevated (Systolic BP 120–129 mmHg and Diastolic BP [DBP] <80 mmHg), Stage 1 Hypertension (SBP 130–139 mmHg or DBP 80–89 mmHg), or Stage 2 Hypertension (SBP ≥140 mmHg or DBP ≥90 mmHg) according to American Heart Association (AHA) guidelines.^8^ When SBP and DBP readings fell into different categories, the more severe category was assigned (e.g., SBP=125 mmHg [Elevated] and DBP=85 mmHg [Stage 1 Hypertension], Classification = Stage 1 Hypertension). Four sensitivity cohorts were defined: the full cohort, which added participants with normal BP (SBP <120 mmHg and DBP <80 mmHg); participants with no recorded antihypertensive medication (no journal entry from 180 days before baseline through the 6-month reading); participants with no recorded GLP-1 receptor agonist use (no journal entry from 30 days before baseline through the 6-month reading); and participants excluding those who first logged antihypertensive medication after baseline.

SBP and DBP trajectories were modeled separately using linear mixed-effects models. Unadjusted models regressed BP on time (baseline, 3-month, 6-month), baseline AHA stratum (Elevated, Stage 1, Stage 2), and their interaction. Adjusted models included core covariates (each interacted with time), season, and measurement time of day as a cyclic term.^31^ 12-month BP was examined with the same models, extending the time factor to 12 months. Sensitivity analyses refit the trajectory models in each of the four sensitivity cohorts, in participants who logged BP at least twice within a week in both the baseline and 6-month windows, and stratified by age (18–39, 40–59, 60–79 years) and sex.

Given that the primary cohort included individuals with Elevated BP or Hypertension, we estimated the effect of regression to the mean (RTM) using a previously described approach.^32^ Expected RTM was calculated as the proportion of baseline BP variance attributable to within-person variability, multiplied by the difference in mean baseline BP between each stratum and the full cohort including participants with normal BP. Because strata were defined jointly by SBP and DBP, the observed stratum mean was used in place of the normal-theory approximation. Within-person variability was estimated as the pooled within-participant variance of BP measurements recorded from baseline through 30 days, among the 14,913 participants (52.8% of the full cohort) contributing two or more measurements in that window. Expected RTM was added to the adjusted model estimate of change to yield an RTM-corrected estimate for each stratum, with 95% confidence intervals from 2,000 participant-level bootstrap resamples. At 12 months, the reference mean and variance were taken from participants contributing 12-month data.

Associations between digital-platform engagement and ≥5 mmHg BP reductions were examined using separate covariate-adjusted logistic regression models. Percentage-based metrics were scaled per 10-percentage-point increment, while average daily app time and total BP reading count were log-transformed due to right-skewed distributions. Each metric was fit in its own covariate-adjusted model and refit in the cohorts with no recorded antihypertensive medication, with no recorded GLP-1 receptor agonist use, and excluding antihypertensive initiation during follow-up.

Associations between continuously measured wearable signals and 6-month BP change were examined for RHR, HRV, physical activity, sleep duration, and sleep consistency. Multivariable logistic regression modeled the odds of achieving a ≥5 mmHg SBP and DBP reduction, with each wearable metric represented by its 30-day pre-baseline mean and 6-month study-window mean. Models adjusted for baseline BP and core covariates and required wearable data on at least 8 of 30 days in the baseline window, ensuring coverage spanned more than a full week (including weekday and weekend days).^33^ Sensitivity analyses refit the model in each of the four sensitivity cohorts. To leverage 6 months of continuous wearable data and every BP reading over the study period, associations were also modeled at the level of the individual readings. For each BP observation, the outcome was deviation from the participant’s baseline BP. Each wearable metric, summarized over a seven-day lookback window, was decomposed into between-person (a participant’s average level) and within-person (fluctuation around that average) components. Models adjusted for core covariates, included a random intercept, and used cluster-robust standard errors on participant, with the cohort restricted to participants contributing at least two non-baseline readings. Sensitivity analyses refit the model in each of the four sensitivity cohorts and, in the primary cohort, using raw BP as the outcome with baseline BP as a covariate.

Analyses used complete cases for all model terms. Linear mixed-effects models, estimated by restricted maximum likelihood (REML), used all available timepoints for each participant. Mixed-effects models carried a participant-level random intercept with Type III tests on Satterthwaite degrees of freedom. Estimated marginal means and contrasts were subjected to within-family Holm correction, with Nakagawa R². Unless otherwise specified, adjusted models included age, sex, baseline body mass index (BMI), and antihypertensive-medication status at baseline. A reduction of ≥5 mmHg in SBP or DBP was prespecified as clinically meaningful.^34^ Logistic coefficients are reported as odds ratios (OR) with 95% confidence intervals (CI). Model diagnostics, including adequacy, collinearity, fit, and calibration were assessed from residual and random-effect distributions, variance inflation factors and, for logistic models, calibration across deciles of predicted probability. Analyses were conducted in R version 4.6.0, with statistical significance defined as α = 0.005.^35^

## Results

### Study Population

The analytic cohort comprised 20,003 adults with a baseline BP classified as Elevated, Stage 1 Hypertension, or Stage 2 Hypertension (Table 1), drawn from 28,259 adults with recorded sex who met the BP-reading criteria.^8^ Participants were predominantly male, and few reported GLP-1 receptor agonist, antihypertensive medication, tobacco, or nicotine use. Mean BMI in each stratum fell within the overweight range (26.8–28.6 kg/m²). Case completeness and subsequent modeled sub-cohorts are described in the supplementary material (**Table S1, Table S2**).

**Table 1:** Characteristics of wearable device users who recorded a blood pressure (BP) classifying them as having Elevated BP, Stage 1 Hypertension, or Stage 2 Hypertension with a follow-up measure 6 months later. BP stratum is defined by current AHA guidelines. Data are presented as mean±SD or n [%].

|  | **Overall<br>(20,003)** | **Elevated<br>(5,163)** | **Stage 1<br>(10,269)** | **Stage 2<br>(4,571)** |
|---|---|---|---|---|
| Age (years) | 45.2±11.3 | 44.4±12.3 | 45.3±10.9 | 45.7±10.7 |
| Sex (M/F) | 18,382/1,621 | 4,810/353 | 9,357/912 | 4,215/356 |
| BMI (kg/m²) | 27.5±4.4 | 26.8±4.0 | 27.4±4.2 | 28.6±4.8 |
| Baseline SBP (mmHg) | 129.6±10.0 | 123.9±2.8 | 127.3±7.0 | 141.4±11.5 |
| Baseline DBP (mmHg) | 82.1±8.4 | 73.7±4.5 | 82.1±4.4 | 91.6±8.5 |
| Study Period (median [IQR]) | 182 [168–202] | 182 [169–202] | 182 [169–202] | 181 [168–200] |
| Tobacco/Nicotine Use (n [%]) | 2,037 [10.2] | 524 [10.1] | 1,036 [10.1] | 477 [10.4] |
| **Antihypertensive Medication Status** |  |  |  |  |
| Established use, ≥8 weeks (n [%]) | 1,498 [7.5] | 354 [6.9] | 758 [7.4] | 386 [8.4] |
| Recent initiation, <8 weeks (n [%]) | 656 [3.3] | 127 [2.5] | 341 [3.3] | 188 [4.1] |
| Initiated during follow-up (n [%]) | 1,528 [7.6] | 313 [6.1] | 706 [6.9] | 509 [11.1] |
| **GLP-1 Receptor Agonist Status** |  |  |  |  |
| Initiated Prior to Study (n [%]) | 241 [1.2] | 60 [1.2] | 146 [1.4] | 35 [0.8] |
| Initiated During Study (n [%]) | 470 [2.3] | 102 [2.0] | 236 [2.3] | 132 [2.9] |

### Blood Pressure Trajectories

Over the approximately 6-month follow-up, both unadjusted and adjusted models demonstrated reductions in SBP across all baseline BP strata (**Table 2**, **Figure 1A).** Participants with Stage 1 and Stage 2 Hypertension experienced reductions in both SBP and DBP by 3 months that were maintained through 6 months, with further reductions in those with Stage 2 Hypertension. SBP was similarly reduced at 3 months and remained lower than baseline at 6 months in those with Elevated BP, while DBP was not different across timepoints. After correction for RTM, adjusted SBP reductions were −2.8 (95% CI −3.2 to −2.3), −3.0 (−3.4 to −2.7) and −6.8 (−7.3 to −6.3) mmHg in participants with Elevated BP, Stage 1 Hypertension, and Stage 2 Hypertension, respectively. DBP changes were −1.2 (−1.5 to −0.9) in Elevated, −2.8 (−3.0 to −2.5) in Stage 1 Hypertension and −5.4 (−5.8 to −5.1) mmHg in Stage 2 Hypertension. Based on a change of ≥5 mmHg,^34^ these BP trajectories resulted in a cohort-wide clinically meaningful SBP reduction of moderate effect size (**Figure 1B**). Overall, 50.2% of users achieved a ≥5 mmHg reduction in SBP, and 27.9% met normal BP by approximately 6 months (**Figure 1C/D**).

**Figure 1:**
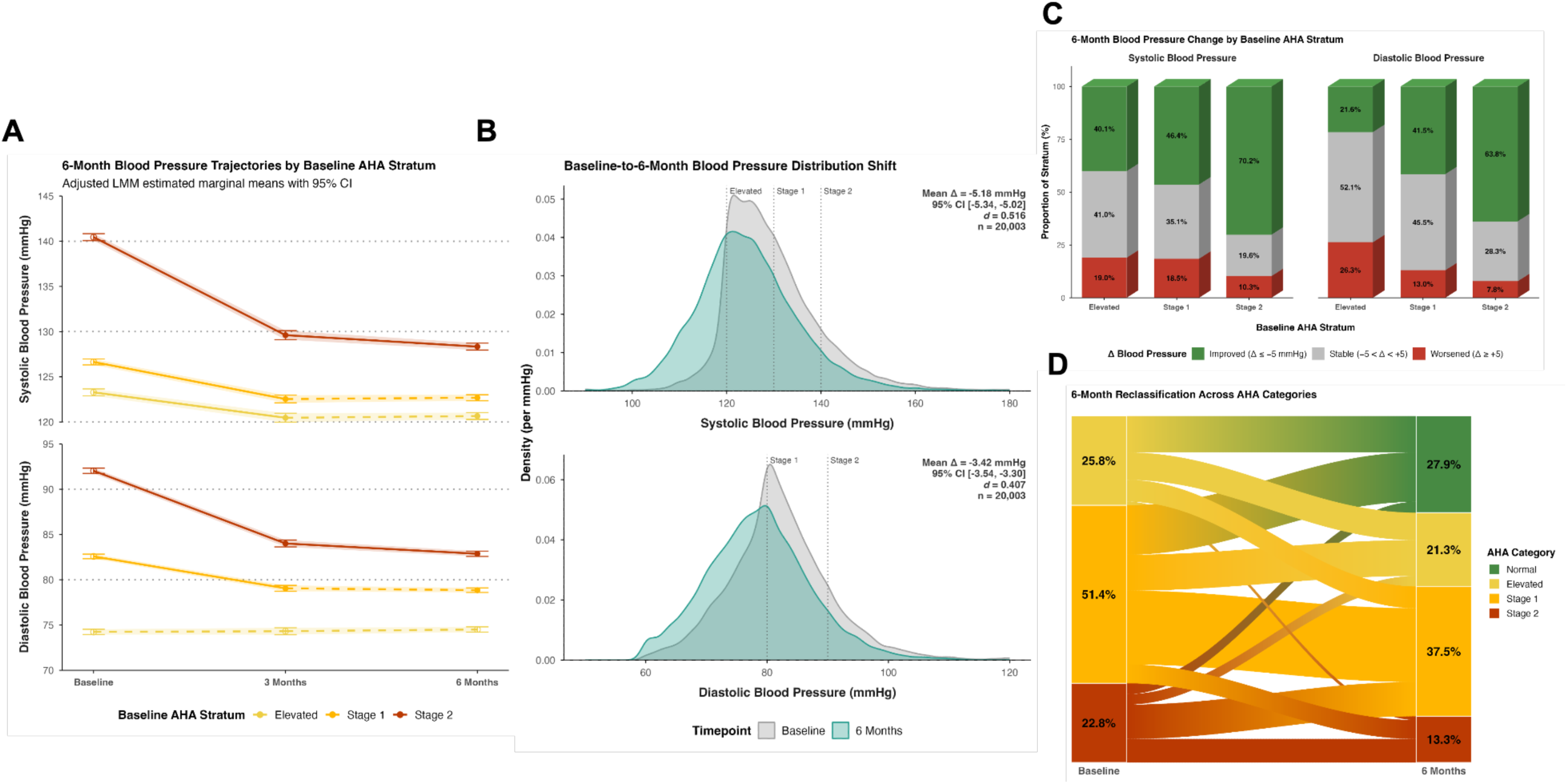
(A) Adjusted mean systolic BP (SBP) and diastolic BP (DBP) at baseline, 3 months, and 6 months by baseline American Heart Association (AHA) stratum (Elevated, Stage 1, Stage 2), from linear mixed models with a participant random intercept; ribbons and error bars are 95% CIs, and dotted lines mark AHA thresholds. Filled points indicate a significant difference from baseline and solid segments a significant change between adjacent timepoints; open points and dashed segments indicate p ≥ 0.005. (B) Kernel-density distributions of SBP (top) and DBP (bottom) at baseline (gray) and 6 months (green); dotted lines mark AHA thresholds. Annotations give the mean change, 95% CI, standardized effect size (d, mean change divided by the pooled baseline SD, as in Table 2), and n. (C) Proportion of each baseline AHA stratum with improved (Δ ≤ −5 mmHg), stable (−5 < Δ < +5 mmHg), or worsened (Δ ≥ +5 mmHg) SBP and DBP at 6 months. (D) Alluvial diagram of movement between AHA categories from baseline to 6 months; band widths are proportional to the share of the cohort following each transition, and percentages are category proportions at each timepoint.

**Table 2:** Blood pressure change from baseline at 3 and 6 months, stratified by baseline AHA stratum. Unadjusted Δ and adjusted Δ are estimated marginal mean changes (mmHg) from baseline, with 95% confidence intervals, from unadjusted and covariate-adjusted linear mixed models with a random intercept per participant. The adjusted model includes age, sex, baseline BMI and baseline antihypertensive medication status, each interacted with time, plus time of day and season. P-values are from the adjusted model and are Holm-corrected within the within-stratum contrast family. Cohen’s d is the absolute adjusted Δ divided by pooled baseline SD of the outcome. Data are presented as EMM (95% CI). R², Nakagawa marginal and conditional R² of the unadjusted and adjusted models; residual skew and excess kurtosis are from the same models.

| Stratum | Time (Months) | Unadjusted Δ (mmHg) | Adjusted Δ (mmHg) | P | *d* |
|---|---|---|---|---|---|
| **Systolic Blood Pressure** |  |  |  |  |  |
| Elevated | 3 | -3.0 (-3.3, -2.6) | -2.8 (-3.3, -2.3) | <0.001 | 0.28 |
|  | 6 | -2.5 (-2.8, -2.2) | -2.6 (-3.1, -2.2) | <0.001 | 0.26 |
| Stage 1 | 3 | -4.1 (-4.4, -3.9) | -4.1 (-4.6, -3.7) | <0.001 | 0.41 |
|  | 6 | -3.7 (-3.9, -3.5) | -4.0 (-4.3, -3.6) | <0.001 | 0.39 |
| Stage 2 | 3 | -10.6 (-11.0, -10.2) | -10.8 (-11.4, -10.3) | <0.001 | 1.08 |
|  | 6 | -11.6 (-11.9, -11.3) | -12.1 (-12.5, -11.7) | <0.001 | 1.21 |
| Marginal R² |  | 0.26 | 0.28 |  |  |
| Conditional R² |  | 0.52 | 0.52 |  |  |
| Residual Skew/Excess Kurtosis |  | 0.23 / 2.3 | 0.23 / 2.4 |  |  |
| **Diastolic Blood Pressure** |  |  |  |  |  |
| Elevated | 3 | 0.3 (0.0, 0.6) | 0.1 (-0.3, 0.5) | 0.700 | 0.01 |
|  | 6 | 0.7 (0.5, 0.9) | 0.3 (-0.1, 0.6) | 0.211 | 0.03 |
| Stage 1 | 3 | -3.2 (-3.4, -3.0) | -3.5 (-3.9, -3.2) | <0.001 | 0.42 |
|  | 6 | -3.2 (-3.4, -3.1) | -3.7 (-4.0, -3.5) | <0.001 | 0.44 |
| Stage 2 | 3 | -7.5 (-7.8, -7.2) | -8.0 (-8.4, -7.6) | <0.001 | 0.96 |
|  | 6 | -8.4 (-8.7, -8.2) | -9.2 (-9.5, -8.8) | <0.001 | 1.09 |
| Marginal R² |  | 0.33 | 0.34 |  |  |
| Conditional R² |  | 0.58 | 0.58 |  |  |
| Residual Skew/Excess Kurtosis |  | 0.22 / 1.6 | 0.22 / 1.6 |  |  |

Sensitivity analyses supported the primary BP findings. Stratification by age and sex revealed direction-concordant changes in BP, and estimates in the full cohort and in the cohorts with no recorded antihypertensive medication, no recorded GLP-1 receptor agonist use, or excluding antihypertensive initiation during follow-up did not substantively differ from the primary cohort (**Table S3**). In the 2,189 participants who recorded BP twice weekly at both baseline and 6 months, adjusted mean reductions were directionally consistent with the primary analysis and retained 54% (Elevated SBP: −1.4 mmHg (95% CI −2.5 to −0.3)) to 83% (Stage 2 Hypertension DBP: −7.6 mmHg (95% CI −8.4 to −6.8)) of the primary adjusted estimates. Among the 1,417 participants (7.1%) contributing 12-month data, BP reductions were at least as large as at 6 months (**Figure S1**), including after correction for RTM (**Table S4**). Participants with Stage 2 Hypertension had further reductions in both SBP and DBP from 6 months, whereas those with Stage 1 Hypertension demonstrated further reductions in SBP while maintaining lower DBP. Participants with Elevated BP maintained lower SBP without significant change in DBP. These improvements resulted in more individuals with Stage 1 and Stage 2 Hypertension at baseline reaching normal BP at follow-up (**Figure S2**).

### Digital Health Platform Engagement

Greater engagement with the wearable-associated digital health platform was consistently associated with higher odds of a ≥5 mmHg SBP and DBP reduction (**Figure 2**). Greater daily app time (SBP: OR = 1.30 [1.25–1.36], p < 0.001; DBP: OR = 1.35 [1.29–1.41], p < 0.001), journal completion (SBP: OR = 1.02 [1.02–1.03], p < 0.001; DBP: OR = 1.02 [1.01–1.03], p < 0.001), notification engagement (SBP: OR = 1.04 [1.03–1.05], p < 0.001; DBP: OR = 1.04 [1.03–1.05], p < 0.001), AI Coach engagement (SBP: OR = 1.07 [1.06–1.09], p < 0.001; DBP: OR = 1.08 [1.06–1.09], p < 0.001), and sleep need achieved (SBP: OR = 1.09 [1.05–1.14], p < 0.001; DBP: OR = 1.16 [1.11–1.21], p < 0.001) were each associated with greater odds of a ≥5 mmHg BP reduction. The proportion of days meeting the platform’s physical activity recommendation was the only engagement metric not associated with BP reduction (SBP: OR = 1.01 [0.98–1.03], p = 0.640; DBP: OR = 1.02 [1.00–1.05], p = 0.079). The number of BP readings recorded was also associated with greater odds of reduction (SBP: OR = 1.28 [1.22–1.33], p < 0.001; DBP: OR = 1.14 [1.09–1.19], p < 0.001). Results were unchanged in the cohorts with no recorded antihypertensive medication, no recorded GLP-1 receptor agonist use, and excluding antihypertensive initiation during follow-up (**Table S5**).

**Figure 2:**
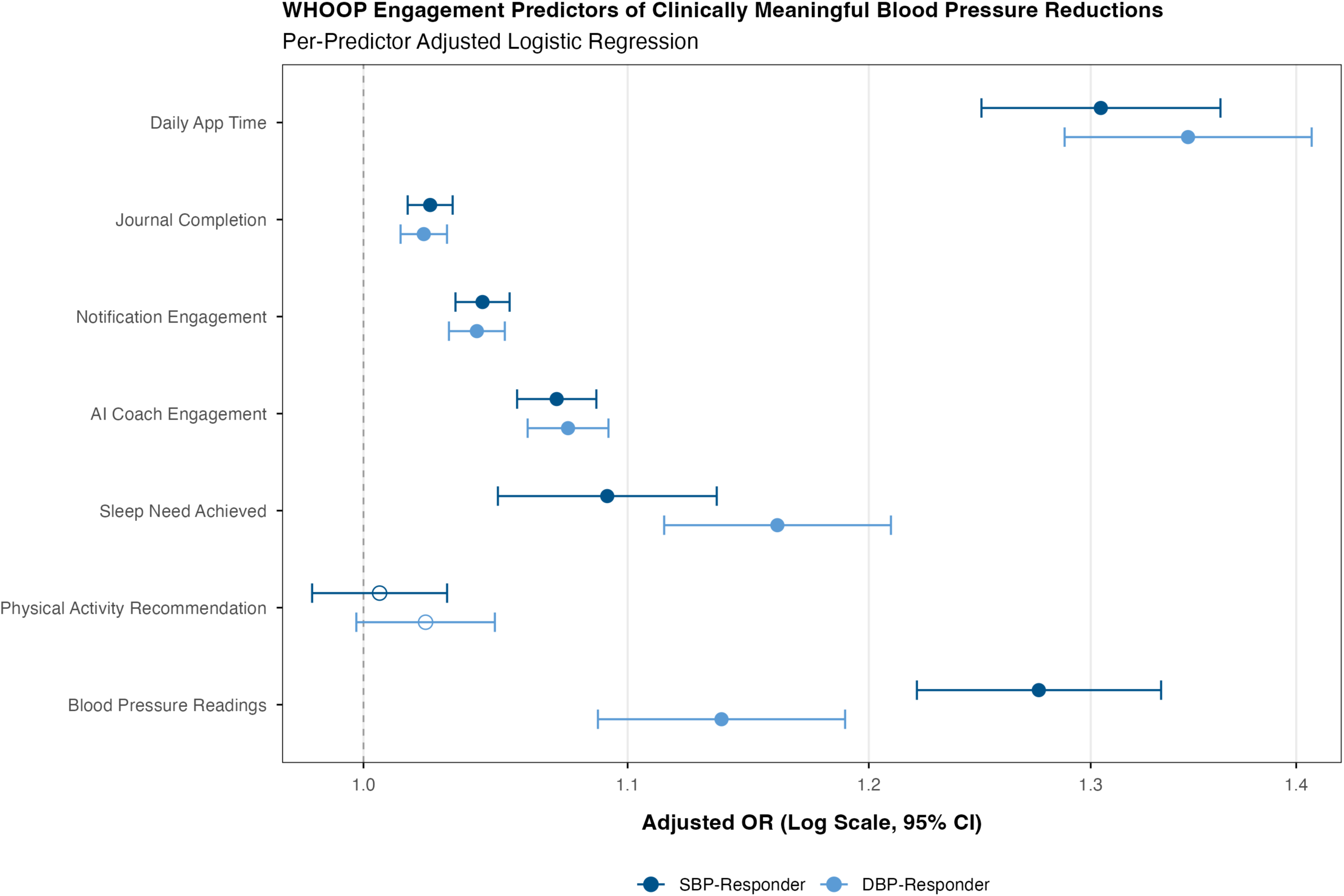
Adjusted odds ratios (ORs) with 95% confidence intervals are shown on a log scale for seven digital health platform engagement metrics: daily in-app time (log-transformed), journal completion, notification engagement, AI Coach engagement, sleep need achieved, physical activity recommendation, and total blood pressure readings taken (log-transformed), for individuals who reduce systolic BP [SBP; blue] and diastolic BP [DBP; light blue], each defined as a ≥5 mmHg reduction from baseline at 6 months. Filled symbols indicate p < 0.005; open symbols indicate p ≥ 0.005. Each predictor is estimated from a separate logistic regression adjusted for the outcome-matched baseline blood pressure, age, sex, BMI, and antihypertensive medication status. Proportion-based predictors are scaled per 10 percentage points; time and reading-count predictors are per log unit.

### Continuous Wearable-Derived Insights

Baseline and follow-up wearable-derived metrics demonstrated numerous associations with ≥5 mmHg BP changes (**Figure 3**). Higher baseline RHR predicted greater odds of meaningful reductions in both SBP and DBP (SBP: OR = 1.026 [1.010–1.042], p = 0.001; DBP: OR = 1.023 [1.007–1.040], p < 0.005), whereas higher RHR during follow-up predicted lower odds of a ≥5 mmHg SBP and DBP reduction (SBP: OR = 0.973 [0.958–0.989], p = 0.001; DBP: OR = 0.966 [0.950–0.982], p < 0.001). Greater physical activity and longer sleep duration during follow-up were associated with greater odds of DBP reduction (physical activity: OR = 1.081 [1.050–1.114], p < 0.001; sleep duration: OR = 1.232 [1.093–1.389], p < 0.001), but not SBP (physical activity: OR = 1.019 [0.990–1.048], p = 0.195; sleep duration: OR = 1.033 [0.921–1.160], p = 0.576). Higher baseline physical activity was associated with lower odds of DBP reduction (OR = 0.950 [0.927–0.974], p < 0.001) but not SBP (OR = 0.986 [0.963–1.010], p = 0.249). These associations were broadly consistent across cohorts (**Table S6**).

**Figure 3:**
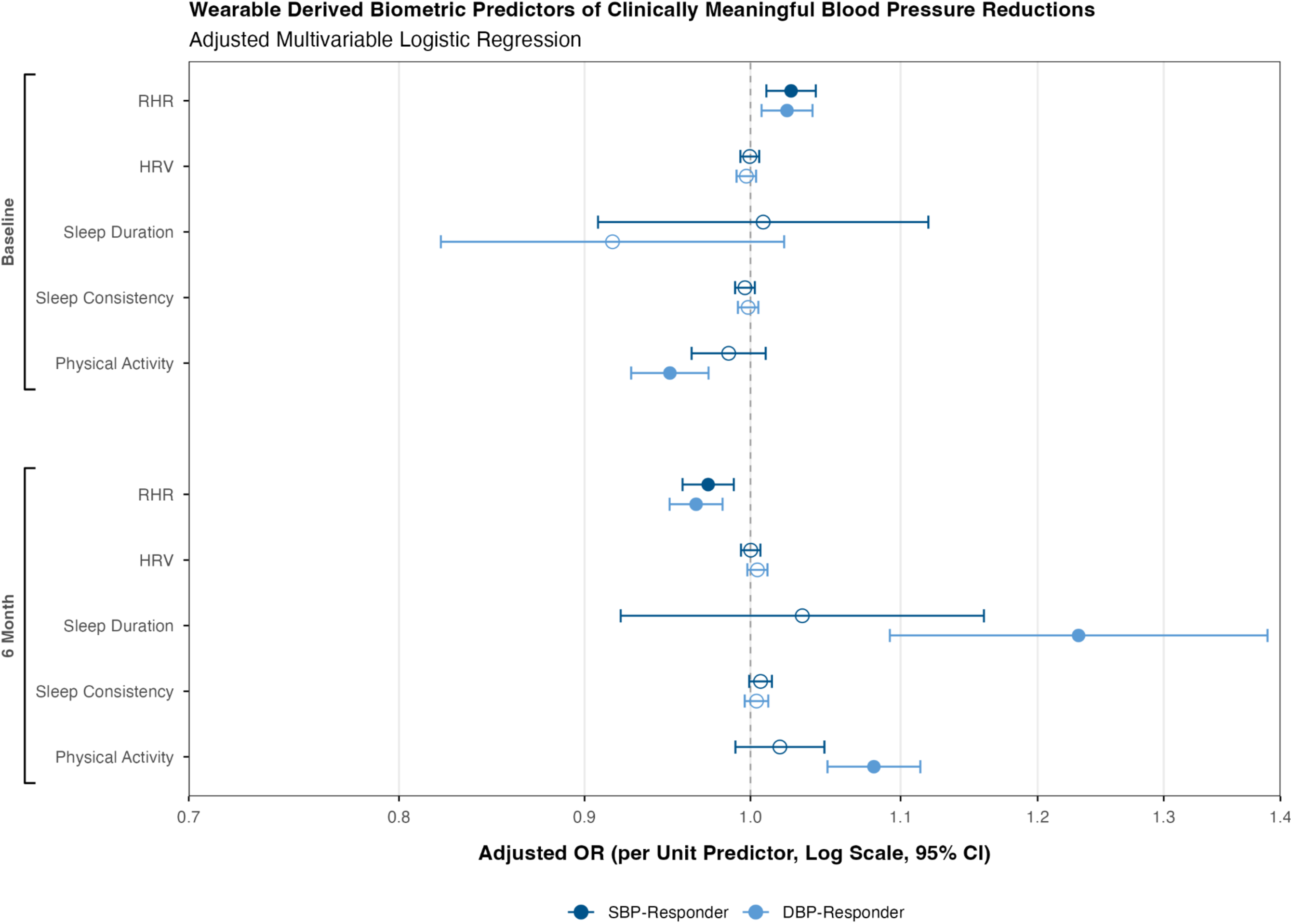
Adjusted odds ratios (ORs) with 95% confidence intervals are shown on a log scale for ten wearable-measured predictors. Each of the five metrics (resting heart rate, heart rate variability, sleep duration, sleep consistency, physical activity) is summarized as a 30-day pre-baseline mean and a 6-month study-period mean for individuals who reduce systolic BP [SBP; blue] and individuals who reduce diastolic BP [DBP; light blue], defined as ≥5 mmHg reduction from baseline at 6 months. Filled symbols indicate p < 0.005; open symbols indicate p ≥ 0.005. All models adjusted for outcome-matched baseline blood pressure, age, sex, BMI, and antihypertensive medication status.

The continuous wearable data throughout follow-up enabled examination of between- and within-person associations between wearable-derived metrics and BP at the reading level (**Figure 4**). At the between-person level, greater habitual physical activity was associated with lower SBP (β = −0.37 mmHg [−0.53 to −0.21], p < 0.001) and DBP (β = −0.41 mmHg [−0.54 to −0.28], p < 0.001), while higher RHR was associated with lower DBP (β = −0.29 mmHg [−0.43 to −0.15], p < 0.001). At the within-person level, acute elevations in RHR were associated with higher SBP (β = 0.56 mmHg [0.47 to 0.65], p < 0.001) and DBP (β = 0.59 mmHg [0.53 to 0.66], p < 0.001), whereas greater-than-usual sleep duration (SBP: β = −0.13 mmHg [−0.19 to −0.08]; DBP: β = −0.07 mmHg [−0.11 to −0.03]; both p < 0.001) and physical activity (SBP: β = −0.18 mmHg [−0.24 to −0.12]; DBP: β = −0.28 mmHg [−0.32 to −0.23]; both p < 0.001) were associated with lower BP. These associations were robust across sensitivity analyses (**Table S7**).

**Figure 4:**
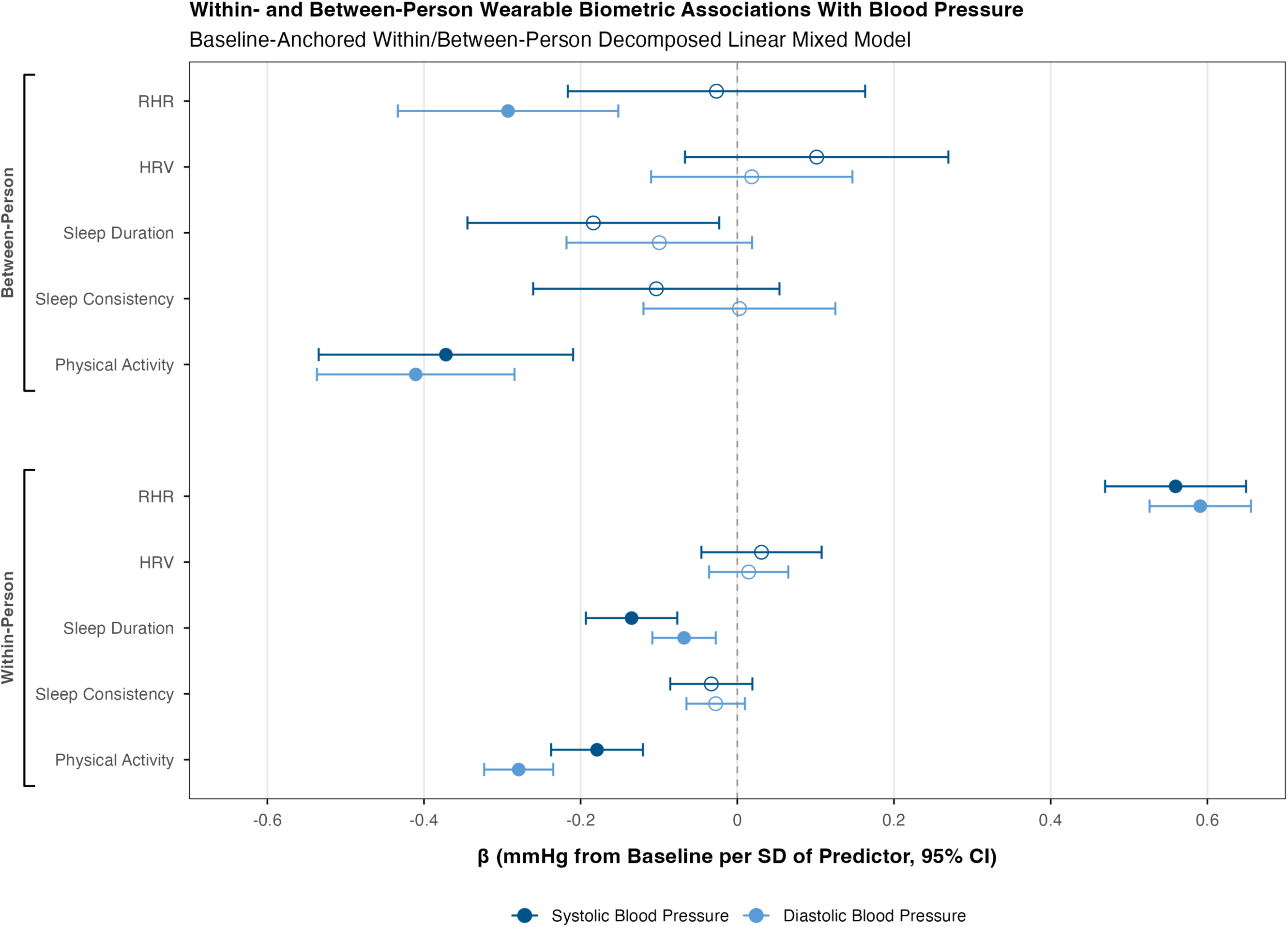
Standardized regression coefficients (β) with 95% confidence intervals are shown on a linear scale for ten wearable-measured daily predictors. Each of the five metrics (resting heart rate, heart rate variability, sleep duration, sleep consistency, physical activity) is decomposed into a between-person component (participant’s mean across all non-baseline readings) and a within-person component (reading-level deviation from that mean) for systolic BP [SBP; blue] and diastolic BP [DBP; light blue], defined as observed BP minus the participant’s own baseline BP at each non-baseline reading. All predictors represent 7-day lookback means standardized to their respective SD; coefficients are interpretable as mmHg of blood pressure change per one SD of the predictor. Filled symbols indicate p < 0.005; open symbols indicate p ≥ 0.005. All models adjusted for age, sex, baseline BMI, and antihypertensive medication status.

## Discussion

We characterized 6-month BP trajectories in adults who wore a consumer wearable device and logged cuff-measured BP in the companion digital health platform. We identified behaviors and biometrics associated with BP change. Across the Elevated and Hypertensive cohort, BP fell over 6 months, with reduction scaling with higher baseline BP. Reductions remained directionally consistent across multiple sensitivity analyses, including analyses using repeated BP measurements and approaches addressing RTM. Greater engagement with the digital platform was associated with higher odds of clinically meaningful BP reduction, while wearable-derived physiology and behavioral metrics were associated with BP reductions both over 6 months and at the individual reading level.

Estimated SBP reductions ranged from 2.6 mmHg in the Elevated stratum to 12.1 mmHg in Stage 2 Hypertension, while DBP reductions ranged from 3.7 mmHg in Stage 1 to 9.2 mmHg in Stage 2. At 12 months, these improvements were sustained or larger. Reductions of this magnitude are likely to be prognostically meaningful, as home-measured BP predicts cardiovascular events independently of, and at least as strongly as, office readings.^10,11^ The magnitude of these reductions aligns with digital hypertension self-management programs, which have reported SBP reductions of up to ∼8 mmHg over 12 months^17^ and 10–20 mmHg over longer follow-up.^14,16^ The proportion of participants achieving a clinically meaningful BP reduction was also similar to that previously reported (∼50%).

When considering scale and follow-up duration, the most comparable data come from a recent analysis of a remote patient care program.^15^ Among 23,638 Medicare enrollees, reductions of 7 mmHg in SBP and 5 mmHg in DBP were observed over ∼30 weeks, with reduction similarly scaling with baseline BP. These reductions were facilitated by a structured clinical program involving a cellular-connected cuff, nurse practitioner-led encounters, and protocolized medication titration. Our findings suggest similar reductions in a setting where the platform itself provided neither clinical contact nor medication management. This is noteworthy because self-monitoring of BP has generally lowered BP only when combined with co-interventions such as feedback, counseling, or medication titration,^13^ and the continuous digital feedback provided by the platform in the present study may be serving an analogous function. Unlike many digital hypertension programs, which depend on documented hypertension, employer or insurer eligibility, or structured coaching,^17,18^ the wearable platform studied here provides BP monitoring and digital guidance without those prerequisites. Taken together, the comparable magnitude of BP reduction observed in this lower-barrier consumer setting suggests that wearable platforms may offer a scalable complement to more resource-intensive hypertension programs. This is consistent with AHA recommendations supporting self-measured and out-of-office BP monitoring,^9,36^ and with the growing adoption of consumer wearables.^19^ Although observational, these findings suggest that RTM-corrected reductions similar in magnitude to those of structured remote programs can occur on a consumer platform that provides no enrollment, eligibility screening, or clinical staffing.

Engagement with the digital health platform assessed by multiple metrics was associated with clinically meaningful BP reduction. Greater time spent in the digital health platform, completion of the in-app journal, engagement with notifications and the AI coach, and adherence to sleep recommendations were each associated with higher odds of a meaningful reduction. The association was particularly evident when engagement was specific to BP, with BP logging among the strongest predictors of clinically meaningful BP reduction. Structured remote care programs have quantified engagement as clinical utilization—vitals transmitted, encounters completed, alerts resolved—and noted that without granular engagement data it is difficult to determine whether engagement itself contributes to BP reduction.^15^ Because the ecosystem studied here captures engagement continuously alongside sleep, physical activity, and autonomic physiology, this association could be examined directly. Given evidence that smartphone application integration with home BP monitoring is not superior to traditional home BP monitoring,^37^ these findings offer promise for wearable ecosystems as a supportive adjunct to BP management. Collectively, our findings support the notion that home and self-measured BP monitoring support BP control,^9,13^ and extend it to a commercially available consumer wearable.

The digital health platform’s continuous behavioral and physiological monitoring allowed us to identify correlates of BP change both across the study period and at the level of individual BP readings. Consistent with established evidence, longer sleep duration and greater physical activity were associated with directionally favorable changes in BP.^36,38^ While dietary changes also plausibly underlie these changes,^6^ the digital health platform did not capture diet and thus precludes evaluation of its relative impact. Wearable-derived physiological metrics also demonstrated associations with BP reductions. Participants achieving BP reduction exhibited favorable RHR trajectories, and at the reading-level, lower BP coincided with periods when an individual’s RHR was lower and their sleep duration and physical activity were higher than typical. These findings extend evidence supporting RHR as a marker of cardiovascular risk^39–41^ by showing that dynamic changes in wearable-derived physiology are associated with BP. More broadly, they demonstrate that consumer wearables can identify both near-term and long-term, person-specific behavioral and physiological changes associated with lower BP. Although these observational findings require prospective validation, they provide proof-of-concept that continuously monitored wearable signals may support personalized hypertension management by identifying and reinforcing behaviors associated with BP improvement.

## Limitations

Several limitations warrant consideration. First, our retrospective real-world observational design cannot establish causality. The trends we observed are associational and cannot rule out reverse causation, such as early BP improvement prompting greater app use. Second, the cohort was restricted to wearable users who recorded an initial BP and returned for another measurement 150–240 days later, likely enriching the sample for individuals who are more health-motivated or health-aware and introducing possible biases (i.e., selection, healthy-user, engagement, and survivorship). The observed BP trajectories therefore characterize this selected group and should not be interpreted as the exclusive effect of initiating BP tracking or wearable use. Generalizability to broader populations, including non-paying adopters, requires prospective study. Relatedly, sex-specific evidence in hypertension remains under-addressed in the literature.^42^ In our stratified analyses, female participants demonstrated BP reductions consistent with the overall study findings, providing direct evidence that the observed pattern was present within the female stratum despite the predominantly male cohort. The underrepresentation of female participants reflects the smaller population meeting the study’s BP-tracking and follow-up criteria and should be considered when interpreting the generalizability of the overall estimates. Further addressing this evidence gap is critical, and future work must specifically interrogate these trends among female individuals, including women and gender-diverse populations. Third, because participants entered the cohort with elevated or hypertensive baseline BP and reductions scaled with baseline severity, RTM is expected to contribute to subsequent reductions.^43,44^ Accounting for RTM attenuated estimates most in the Stage 2 stratum, indicating that the scaling of reduction with baseline severity is partly attributable to RTM, but reductions nonetheless remained across strata. Notably, mean baseline SBP in the Elevated stratum was roughly equivalent to the full cohort mean, such that expected RTM was negligible (0.1 mmHg), yet a reduction was still observed. Sensitivity analyses using repeated BP measurements attenuated but did not eliminate the observed changes, supporting persistence of the overall pattern. Fourth, our metric of physical activity is primarily derived from intra-day heart rate fluctuations, which may underestimate activities with primarily musculoskeletal demands (e.g., resistance training). Finally, BP measurements and several behavioral variables relied on participant-reported data and are subject to measurement, recall, and reporting bias.^45^ Medication and tobacco/nicotine exposure depended on voluntary in-app reporting. Absence of a recorded medication entry cannot establish medication nonuse, and residual confounding from pharmacologic treatment is possible.

## Conclusions

To our knowledge, this is the first report of BP trajectories among individuals using a commercially available consumer wearable and companion digital health platform outside of a structured hypertension management program. Greater engagement with the digital health platform was consistently associated with clinically meaningful BP reduction. Additionally, continuous wearable data identified behavioral and physiological states associated with both short-term BP responses and longer-term BP improvements. Together, these findings highlight the potential of consumer wearables as scalable platforms for supporting BP monitoring and management in real-world settings.

## Author Contributions

CPC, and GJG conceived the study with contributions from JGC, ABB, WvH, FF, LRP, DC, CJC, and KEH. CPC led the methodology. CPC and GJG led data curation and formal analysis. CPC prepared all figures, with input from GJG and JGC. CPC drafted the manuscript. All authors reviewed and edited the manuscript and approved the final version.

## Acknowledgements

The authors wish to acknowledge the technical assistance of the WHOOP Machine Learning and Research and Data Engineering teams for their contributions to feature development and data infrastructure. We also thank the broader WHOOP employee community for their continued support of the company’s research initiatives.

## Code Availability

The analysis code that supports the findings of this study is publicly available at https://github.com/christiancheung-w/bp-cohort-analysis

## Data Availability

The participant data are not publicly available due to intellectual property concerns of WHOOP, Inc. Deidentified participant data and a data dictionary may be made available upon reasonable request to WHOOP (via). Access requires submission of a methodologically sound proposal, approved by WHOOP’s research team, and a signed data use agreement. Requests will receive a response within 4 weeks.

## AI Diligence Statement

The authors used the generative AI tool Claude (Anthropic) for language editing and code review. After using this tool, the authors reviewed and edited the content as needed and take full responsibility for the content of the published article.

---

**Figure S1:** Adjusted mean systolic BP (SBP) and diastolic BP (DBP) at baseline, 3, 6, and 12 months by baseline American Heart Association (AHA) stratum (Elevated, Stage 1, Stage 2), from linear mixed models with a participant random intercept; ribbons and error bars are 95% CIs, and dotted lines mark AHA thresholds. Filled points indicate a significant difference from baseline and solid segments a significant change between adjacent timepoints; open points and dashed segments indicate p ≥ 0.005.

**Figure S2:** Gradient alluvial diagram representing individual movement between AHA blood pressure categories from baseline to 6 and 12 months among the 1,417 participants with 12-month data; band widths are proportional to the share of these participants following each transition, and percentages represent the category proportions at each timepoint.

