## Supplementary Information for "Real-World Cuff-Measured Blood Pressure Trajectories in 20,003 Consumer Wearable Users"

**Contents**

Figure S1. Adjusted mean systolic and diastolic blood pressure at baseline, 3, 6, and 12 months by baseline AHA stratum

Figure S2. Movement between AHA blood pressure categories from baseline to 6 and 12 months among participants with 12-month data

Table S1. Case completeness counts and rates for all variables

Table S2. Participants and observations contributing to each model

Table S3. Six-month change in blood pressure from baseline in the primary cohort and in sensitivity cohorts and subgroups, by baseline blood pressure category

Table S4. Blood pressure change from baseline at 3, 6 and 12 months by baseline blood pressure category, from the model including the 12-month timepoint

Table S5. Associations of platform engagement with a ≥5 mmHg reduction in blood pressure at 6 months in the primary cohort and in sensitivity cohorts

Table S6. Associations of baseline and 6-month wearable-derived metrics with a ≥5 mmHg reduction in blood pressure at 6 months in the primary cohort and in sensitivity cohorts

Table S7. Between- and within-person associations of wearable-derived metrics with blood pressure at the reading level in the primary cohort and in sensitivity analyses

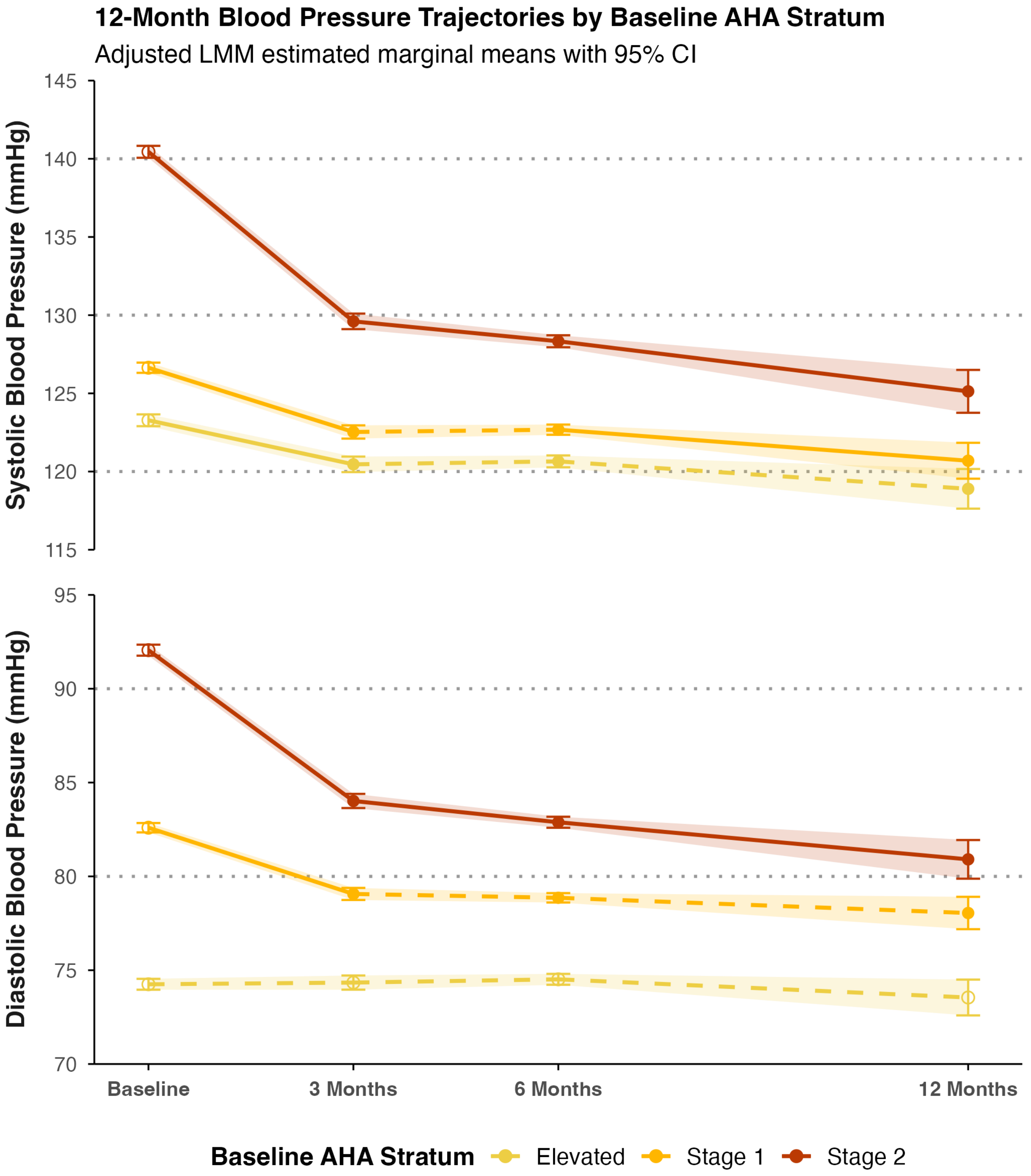

**Figure S1:** Adjusted mean systolic BP (SBP) and diastolic BP (DBP) at baseline, 3, 6, and 12 months by baseline American Heart Association (AHA) stratum (Elevated, Stage 1, Stage 2), from linear mixed models with a participant random intercept; ribbons and error bars are 95% CIs, and dotted lines mark AHA thresholds. Filled points indicate a significant difference from baseline and solid segments a significant change between adjacent timepoints; open points and dashed segments indicate $p \geq 0.005$.

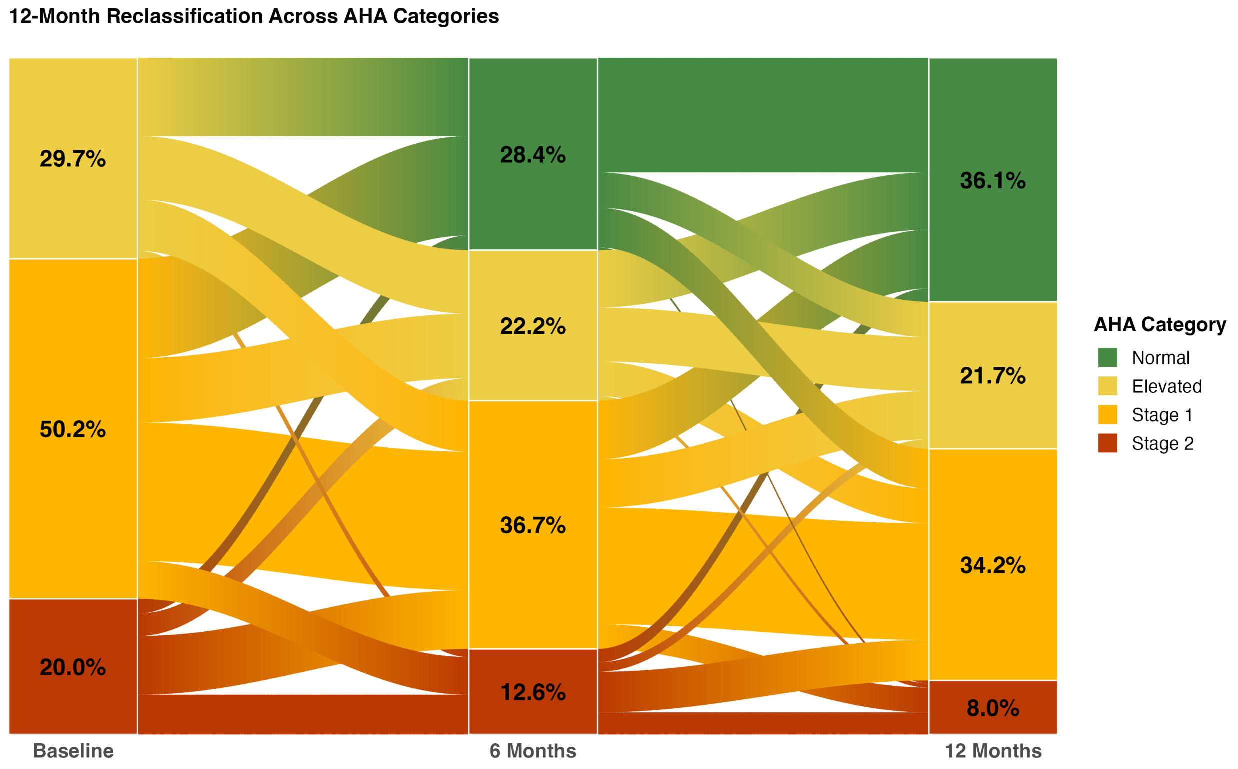

**Figure S2:** Gradient alluvial diagram representing individual movement between AHA blood pressure categories from baseline to 6 and 12 months among the 1,417 participants with 12-month data; band widths are proportional to the share of these participants following each transition, and percentages represent the category proportions at each timepoint.

| User-Level (N = 20,003) | | |
|---|---|---|
| **Variable** | **n** | **% of Users** |
| **Blood Pressure** | | |
| 3-Month Blood Pressure | 10,246 | 51.2 |
| 6-Month Blood Pressure | 20,003 | 100.0 |
| 12-Month Blood Pressure | 1,417 | 7.1 |
| **Core Covariates** | | |
| Age | 20,003 | 100.0 |
| Sex | 20,003 | 100.0 |
| Baseline BMI | 19,986 | 99.9 |
| **Wearable Biometrics** | | |
| Baseline RHR | 14,230 | 71.1 |
| 3-Month RHR | 19,634 | 98.2 |
| 6-Month RHR | 19,262 | 96.3 |
| Baseline HRV | 14,229 | 71.1 |
| 3-Month HRV | 19,631 | 98.1 |
| 6-Month HRV | 19,259 | 96.3 |
| Baseline Physical Activity | 14,230 | 71.1 |
| 3-Month Physical Activity | 19,634 | 98.2 |
| 6-Month Physical Activity | 19,262 | 96.3 |
| Baseline Sleep Duration | 14,230 | 71.1 |
| 3-Month Sleep Duration | 19,634 | 98.2 |
| 6-Month Sleep Duration | 19,262 | 96.3 |
| Baseline Sleep Consistency | 14,229 | 71.1 |
| 3-Month Sleep Consistency | 19,634 | 98.2 |
| 6-Month Sleep Consistency | 19,262 | 96.3 |
| **Engagement Predictors** | | |
| Days Journaled | 20,003 | 100.0 |
| Average Daily App Time | 19,996 | 100.0 |
| Days Engaged with Notifications | 20,003 | 100.0 |
| Days Used AI Coach | 20,003 | 100.0 |
| Days Fulfilled Sleep Need | 19,997 | 100.0 |
| Days Met Physical Activity Recommendation | 19,967 | 99.8 |
| Number of Blood Pressure Readings | 20,003 | 100.0 |

| Reading Level (N = 208,356) | | |
|---|---|---|
| **Variable** | **n** | **% of Readings** |
| RHR | 195,764 | 94.0 |
| HRV | 195,733 | 93.9 |
| Physical Activity | 195,768 | 94.0 |
| Sleep Duration | 195,761 | 94.0 |
| Sleep Consistency | 192,245 | 92.3 |

**Table S1:** Case completeness counts and rates for all variables.

**Table S2. Participants and observations contributing to each model**

| Analysis | Cohort | Outcome | Predictor set | Participants (n) | Observations (n) | Events (SBP / DBP) | Cohort ceiling (n) | % of ceiling | Fit / diagnostics |
|---|---|---|---|---|---|---|---|---|---|
| **Primary analyses** | | | | | | | | | |
| 6-month BP trajectory (LMM) | Primary cohort | - | Adjusted | 19,986 | 50,209 | - | 20,003 | 99.9 | $R^2$ 0.28 / 0.52 (SBP), 0.34 / 0.58 (DBP) |
| 6-month BP trajectory (LMM) | Primary cohort | - | Unadjusted | 20,003 | 50,252 | - | 20,003 | 100.0 | $R^2$ 0.26 / 0.52 (SBP), 0.33 / 0.58 (DBP) |
| 12-month BP trajectory (LMM, 4 timepoints) | Primary cohort | - | Adjusted | 19,986 | 51,626 | - | 20,003 | 99.9 | $R^2$ 0.28 / 0.53 (SBP), 0.34 / 0.58 (DBP) |
| 12-month BP trajectory (LMM, 4 timepoints) | Primary cohort | - | Unadjusted | 20,003 | 51,669 | - | 20,003 | 100.0 | $R^2$ 0.25 / 0.52 (SBP), 0.33 / 0.58 (DBP) |
| Wearable metrics (logistic) | Primary cohort | ≥5 mmHg reduction | Primary | 14,189 | 14,189 | 7,051 / 5,702 | 14,230 | 99.7 | Calibration ≤2.3 / 2.9 pp; VIF ≤15.6 |
| Reading-level within/between-person model (LMM) | Primary cohort | - | Anchored | 16,161 | 132,843 | - | 20,003 | 80.8 | $R^2$ 0.007 / 0.58 (SBP), 0.012 / 0.61 (DBP) |
| Engagement (logistic, separate models) | Primary cohort | ≥5 mmHg reduction | Sleep need achieved | 19,980 | 19,980 | 10,026 / 8,280 | 20,003 | 99.9 | Calibration ≤1.8 / 1.8 pp; VIF 1.10 |
| Engagement (logistic, separate models) | Primary cohort | ≥5 mmHg reduction | Daily app time | 19,979 | 19,979 | 10,026 / 8,282 | 20,003 | 99.9 | Calibration ≤1.9 / 1.9 pp; VIF 1.70 |
| Engagement (logistic, separate models) | Primary cohort | ≥5 mmHg reduction | Blood pressure readings | 19,986 | 19,986 | 10,029 / 8,284 | 20,003 | 99.9 | Calibration ≤2.8 / 2.0 pp; VIF 1.12 |
| Engagement (logistic, separate models) | Primary cohort | ≥5 mmHg reduction | Notification engagement | 19,986 | 19,986 | 10,029 / 8,284 | 20,003 | 99.9 | Calibration ≤2.2 / 1.2 pp; VIF 1.57 |
| Engagement (logistic, separate models) | Primary cohort | ≥5 mmHg reduction | Journal completion | 19,986 | 19,986 | 10,029 / 8,284 | 20,003 | 99.9 | Calibration ≤1.6 / 2.0 pp; VIF 1.26 |
| Engagement (logistic, separate models) | Primary cohort | ≥5 mmHg reduction | Physical activity recommendation | 19,950 | 19,950 | 10,009 / 8,263 | 20,003 | 99.7 | Calibration ≤1.5 / 1.7 pp; VIF 1.04 |
| Engagement (logistic, separate models) | Primary cohort | ≥5 mmHg reduction | AI Coach engagement | 19,986 | 19,986 | 10,029 / 8,284 | 20,003 | 99.9 | Calibration ≤1.6 / 1.4 pp; VIF 1.71 |
| **Sensitivity analyses** | | | | | | | | | |
| 6-month BP trajectory (LMM) | Twice-weekly readings | - | Adjusted | 2,189 | 5,977 | - | 2,190 | 100.0 | $R^2$ 0.29 / 0.58 (SBP), 0.35 / 0.67 (DBP) |
| 6-month BP trajectory (LMM) | Twice-weekly readings | - | Unadjusted | 2,190 | 5,980 | - | 2,190 | 100.0 | $R^2$ 0.27 / 0.58 (SBP), 0.35 / 0.67 (DBP) |
| 6-month BP trajectory (LMM) | Age 18–39 | - | Adjusted | 6,542 | 15,782 | - | 6,546 | 99.9 | — |
| 6-month BP trajectory (LMM) | Age 40–59 | - | Adjusted | 11,244 | 28,650 | - | 11,256 | 99.9 | — |
| 6-month BP trajectory (LMM) | Age 60–79 | - | Adjusted | 2,200 | 5,777 | - | 2,201 | 100.0 | — |
| 6-month BP trajectory (LMM) | Female | - | Adjusted | 1,619 | 4,008 | - | 1,621 | 99.9 | — |
| 6-month BP trajectory (LMM) | Male | - | Adjusted | 18,367 | 46,201 | - | 18,382 | 99.9 | — |
| 6-month BP trajectory (LMM) | Full cohort (including normal BP) | - | Adjusted | 28,238 | 70,717 | - | 28,259 | 99.9 | $R^2$ 0.44 / 0.63 (SBP), 0.41 / 0.63 (DBP) |
| 6-month BP trajectory (LMM) | Full cohort (including normal BP) | - | Unadjusted | 28,259 | 70,770 | - | 28,259 | 100.0 | $R^2$ 0.41 / 0.62 (SBP), 0.40 / 0.63 (DBP) |
| 6-month BP trajectory (LMM) | No recorded antihypertensive medication | - | Adjusted | 16,305 | 40,546 | - | 16,321 | 99.9 | $R^2$ 0.28 / 0.52 (SBP), 0.34 / 0.58 (DBP) |
| 6-month BP trajectory (LMM) | No recorded antihypertensive medication | - | Unadjusted | 16,321 | 40,586 | - | 16,321 | 100.0 | $R^2$ 0.25 / 0.51 (SBP), 0.33 / 0.58 (DBP) |

| Analysis | Cohort | Outcome | Predictor set | Participants (n) | Observations (n) | Events (SBP / DBP) | Cohort ceiling (n) | % of ceiling | Fit / diagnostics |
|---|---|---|---|---|---|---|---|---|---|
| 6-month BP trajectory (LMM) | No recorded GLP-1 receptor agonist use | - | Adjusted | 19,275 | 48,380 | - | 19,292 | 99.9 | — |
| 6-month BP trajectory (LMM) | No recorded GLP-1 receptor agonist use | - | Unadjusted | 19,292 | 48,423 | - | 19,292 | 100.0 | — |
| 6-month BP trajectory (LMM) | Excluding antihypertensive initiation during follow-up | - | Adjusted | 18,458 | 46,160 | - | 18,475 | 99.9 | — |
| 6-month BP trajectory (LMM) | Excluding antihypertensive initiation during follow-up | - | Unadjusted | 18,475 | 46,203 | - | 18,475 | 100.0 | — |
| RTM correction, 6 months (Barnett) | Primary cohort | - | RTM-adjusted change | 20,003 | 20,003 | - | 20,003 | 100.0 | — |
| RTM correction, 12 months (Barnett) | 12-month completers | - | RTM-adjusted change | 1,417 | 1,417 | - | 1,417 | 100.0 | — |
| Wearable metrics (logistic) | Full cohort (including normal BP) | ≥5 mmHg reduction | Primary | 20,360 | 20,360 | 8,214 / 6,809 | 20,416 | 99.7 | — |
| Wearable metrics (logistic) | No recorded antihypertensive medication | ≥5 mmHg reduction | Primary | 11,831 | 11,831 | 5,914 / 4,781 | 11,865 | 99.7 | — |
| Wearable metrics (logistic) | No recorded GLP-1 receptor agonist use | ≥5 mmHg reduction | Primary | 13,698 | 13,698 | 6,779 / 5,466 | 13,738 | 99.7 | — |
| Wearable metrics (logistic) | Excluding antihypertensive initiation during follow-up | ≥5 mmHg reduction | Primary | 13,751 | 13,751 | 6,808 / 5,504 | 13,791 | 99.7 | — |
| Reading-level within/between-person model (LMM) | Full cohort (including normal BP) | - | Anchored | 22,408 | 185,310 | - | 28,259 | 79.3 | — |
| Reading-level within/between-person model (LMM) | No recorded antihypertensive medication | - | Anchored | 12,893 | 97,586 | - | 16,321 | 79.0 | — |
| Reading-level within/between-person model (LMM) | No recorded GLP-1 receptor agonist use | - | Anchored | 15,543 | 126,955 | - | 19,292 | 80.6 | — |
| Reading-level within/between-person model (LMM) | Excluding antihypertensive initiation during follow-up | - | Anchored | 14,765 | 116,813 | - | 18,475 | 79.9 | — |
| Reading-level model, raw BP outcome (LMM) | Primary cohort | - | Anchored | 16,161 | 132,843 | - | 20,003 | 80.8 | — |
| Engagement (logistic, separate models) | No recorded antihypertensive medication | ≥5 mmHg reduction | Sleep need achieved | 16,300 | 16,300 | 8,182 / 6,765 | 16,321 | 99.9 | Calibration ≤2.2 / 2.8 pp |
| Engagement (logistic, separate models) | No recorded antihypertensive medication | ≥5 mmHg reduction | Daily app time | 16,299 | 16,299 | 8,181 / 6,767 | 16,321 | 99.9 | Calibration ≤2.3 / 2.1 pp |
| Engagement (logistic, separate models) | No recorded antihypertensive medication | ≥5 mmHg reduction | Blood pressure readings | 16,305 | 16,305 | 8,184 / 6,769 | 16,321 | 99.9 | Calibration ≤2.9 / 1.9 pp |

| Analysis | Cohort | Outcome | Predictor set | Participants (n) | Observations (n) | Events (SBP / DBP) | Cohort ceiling (n) | % of ceiling | Fit / diagnostics |
|---|---|---|---|---|---|---|---|---|---|
| Engagement (logistic, separate models) | No recorded antihypertensive medication | ≥5 mmHg reduction | Notification engagement | 16,305 | 16,305 | 8,184 / 6,769 | 16,321 | 99.9 | Calibration ≤2.3 / 2.6 pp |
| Engagement (logistic, separate models) | No recorded antihypertensive medication | ≥5 mmHg reduction | Journal completion | 16,305 | 16,305 | 8,184 / 6,769 | 16,321 | 99.9 | Calibration ≤2.0 / 2.1 pp |
| Engagement (logistic, separate models) | No recorded antihypertensive medication | ≥5 mmHg reduction | Physical activity recommendation | 16,274 | 16,274 | 8,166 / 6,750 | 16,321 | 99.7 | Calibration ≤2.4 / 1.8 pp |
| Engagement (logistic, separate models) | No recorded antihypertensive medication | ≥5 mmHg reduction | AI Coach engagement | 16,305 | 16,305 | 8,184 / 6,769 | 16,321 | 99.9 | Calibration ≤2.3 / 1.7 pp |
| Engagement (logistic, separate models) | No recorded GLP-1 receptor agonist use | ≥5 mmHg reduction | Sleep need achieved | 19,269 | 19,269 | 9,617 / 7,923 | 19,292 | 99.9 | — |
| Engagement (logistic, separate models) | No recorded GLP-1 receptor agonist use | ≥5 mmHg reduction | Daily app time | 19,269 | 19,269 | 9,617 / 7,925 | 19,292 | 99.9 | — |
| Engagement (logistic, separate models) | No recorded GLP-1 receptor agonist use | ≥5 mmHg reduction | Blood pressure readings | 19,275 | 19,275 | 9,620 / 7,927 | 19,292 | 99.9 | — |
| Engagement (logistic, separate models) | No recorded GLP-1 receptor agonist use | ≥5 mmHg reduction | Notification engagement | 19,275 | 19,275 | 9,620 / 7,927 | 19,292 | 99.9 | — |
| Engagement (logistic, separate models) | No recorded GLP-1 receptor agonist use | ≥5 mmHg reduction | Journal completion | 19,275 | 19,275 | 9,620 / 7,927 | 19,292 | 99.9 | — |
| Engagement (logistic, separate models) | No recorded GLP-1 receptor agonist use | ≥5 mmHg reduction | Physical activity recommendation | 19,240 | 19,240 | 9,600 / 7,906 | 19,292 | 99.7 | — |
| Engagement (logistic, separate models) | No recorded GLP-1 receptor agonist use | ≥5 mmHg reduction | AI Coach engagement | 19,275 | 19,275 | 9,620 / 7,927 | 19,292 | 99.9 | — |
| Engagement (logistic, separate models) | Excluding antihypertensive initiation during follow-up | ≥5 mmHg reduction | Sleep need achieved | 18,453 | 18,453 | 9,197 / 7,601 | 18,475 | 99.9 | — |
| Engagement (logistic, separate models) | Excluding antihypertensive initiation during follow-up | ≥5 mmHg reduction | Daily app time | 18,451 | 18,451 | 9,196 / 7,603 | 18,475 | 99.9 | — |
| Engagement (logistic, separate models) | Excluding antihypertensive initiation during follow-up | ≥5 mmHg reduction | Blood pressure readings | 18,458 | 18,458 | 9,199 / 7,605 | 18,475 | 99.9 | — |
| Engagement (logistic, separate models) | Excluding antihypertensive initiation during follow-up | ≥5 mmHg reduction | Notification engagement | 18,458 | 18,458 | 9,199 / 7,605 | 18,475 | 99.9 | — |
| Engagement (logistic, separate models) | Excluding antihypertensive initiation during follow-up | ≥5 mmHg reduction | Journal completion | 18,458 | 18,458 | 9,199 / 7,605 | 18,475 | 99.9 | — |
| Engagement (logistic, separate models) | Excluding antihypertensive initiation during follow-up | ≥5 mmHg reduction | Physical activity recommendation | 18,424 | 18,424 | 9,180 / 7,585 | 18,475 | 99.7 | — |

| Analysis | Cohort | Outcome | Predictor set | Participants (n) | Observations (n) | Events (SBP / DBP) | Cohort ceiling (n) | % of ceiling | Fit / diagnostics |
|---|---|---|---|---|---|---|---|---|---|
| Engagement (logistic, separate models) | Excluding antihypertensive initiation during follow-up | ≥5 mmHg reduction | AI Coach engagement | 18,458 | 18,458 | 9,199 / 7,605 | 18,475 | 99.9 | — |

Participants, distinct participants in the fitted model. Observations, rows in the fitted model (participant-timepoints for trajectory models; BP readings for reading-level models; participants for logistic models). Events, participants with a ≥5 mmHg reduction in SBP / DBP at 6 months. Twice-weekly readings, participants with at least two readings within a week in both the baseline and 6-month windows. Cohort ceiling, participants eligible for that analysis; % of ceiling = participants / cohort ceiling. Wearable-metric models require ≥8 days of wear in the baseline window, so their ceilings are smaller. The 12-month RTM correction is restricted to participants with a 12-month reading (1,417 of 20,003). No recorded antihypertensive medication: no journal entry from 180 days before baseline through the 6-month reading. No recorded GLP-1 receptor agonist use: no journal entry from 30 days before baseline through the 6-month reading. Excluding antihypertensive initiation during follow-up: excludes participants whose first logged antihypertensive medication was after baseline. BP, blood pressure; DBP, diastolic blood pressure; GLP-1, glucagon-like peptide-1; LMM, linear mixed-effects model; RTM, regression to the mean; SBP, systolic blood pressure. Fit / diagnostics: $R^2$, Nakagawa marginal / conditional $R^2$ of the fitted model; for logistic models, the largest absolute difference between observed and predicted proportions across deciles of predicted probability (SBP / DBP) and the largest variance inflation factor (VIF; engagement metrics: from the model with all seven metrics entered jointly); —, not computed.

**Table S3. Six-month change in blood pressure from baseline in the primary cohort and in sensitivity cohorts and subgroups, by baseline blood pressure category**

| Analysis | n | SBP change, mmHg (95% CI) | DBP change, mmHg (95% CI) |
|---|---|---|---|
| **Elevated BP** | | | |
| Primary cohort | 5,163 | −2.6 (−3.1, −2.2) | 0.3 (−0.1, 0.6) |
| Full cohort (including normal BP) | 5,163 | −2.9 (−3.3, −2.6) | 0.1 (−0.1, 0.4) |
| No recorded antihypertensive medication | 4,369 | −3.4 (−3.8, −3.0) | −0.2 (−0.5, 0.1) |
| No recorded GLP-1 receptor agonist use | 5,001 | −2.5 (−3.0, −2.1) | 0.3 (0.0, 0.7) |
| Excluding antihypertensive initiation during follow-up | 4,850 | −2.7 (−3.2, −2.3) | 0.2 (−0.1, 0.5) |
| Female | 353 | −3.4 (−4.9, −2.0) | 0.0 (−1.1, 1.0) |
| Male | 4,810 | −2.1 (−2.5, −1.8) | 0.6 (0.4, 0.9) |
| Age 18–39 years | 1,992 | −2.3 (−3.1, −1.5) | 0.2 (−0.4, 0.9) |
| Age 40–59 years | 2,485 | −2.5 (−3.1, −1.9) | 0.5 (0.0, 0.9) |
| Age 60–79 years | 686 | −3.4 (−4.5, −2.3) | −0.4 (−1.1, 0.3) |
| **Stage 1 Hypertension** | | | |
| Primary cohort | 10,258 | −4.0 (−4.3, −3.6) | −3.7 (−4.0, −3.5) |
| Full cohort (including normal BP) | 10,258 | −4.3 (−4.6, −4.0) | −3.9 (−4.1, −3.6) |
| No recorded antihypertensive medication | 8,453 | −4.7 (−5.0, −4.3) | −4.2 (−4.5, −3.9) |
| No recorded GLP-1 receptor agonist use | 9,876 | −3.8 (−4.2, −3.5) | −3.6 (−3.9, −3.3) |
| Excluding antihypertensive initiation during follow-up | 9,552 | −4.1 (−4.5, −3.7) | −3.8 (−4.1, −3.6) |
| Female | 910 | −4.0 (−5.1, −2.9) | −4.3 (−5.0, −3.5) |
| Male | 9,348 | −3.6 (−3.9, −3.2) | −3.3 (−3.6, −3.1) |
| Age 18–39 years | 3,217 | −3.7 (−4.4, −3.0) | −4.0 (−4.6, −3.4) |
| Age 40–59 years | 5,965 | −3.8 (−4.2, −3.3) | −3.5 (−3.9, −3.2) |
| Age 60–79 years | 1,076 | −5.2 (−6.2, −4.2) | −3.9 (−4.5, −3.2) |
| **Stage 2 Hypertension** | | | |
| Primary cohort | 4,565 | −12.1 (−12.5, −11.7) | −9.2 (−9.5, −8.8) |
| Full cohort (including normal BP) | 4,565 | −12.5 (−12.9, −12.1) | −9.3 (−9.6, −9.0) |
| No recorded antihypertensive medication | 3,483 | −12.6 (−13.0, −12.1) | −9.6 (−9.9, −9.2) |
| No recorded GLP-1 receptor agonist use | 4,398 | −11.9 (−12.3, −11.4) | −9.0 (−9.3, −8.7) |
| Excluding antihypertensive initiation during follow-up | 4,056 | −12.0 (−12.4, −11.5) | −9.1 (−9.5, −8.8) |
| Female | 356 | −12.6 (−14.0, −11.2) | −9.7 (−10.7, −8.7) |
| Male | 4,209 | −11.7 (−12.1, −11.3) | −8.8 (−9.1, −8.5) |
| Age 18–39 years | 1,333 | −11.9 (−12.7, −11.0) | −9.9 (−10.5, −9.2) |
| Age 40–59 years | 2,794 | −11.7 (−12.3, −11.1) | −8.9 (−9.4, −8.5) |
| Age 60–79 years | 438 | −14.8 (−16.1, −13.6) | −8.3 (−9.1, −7.4) |

Values are estimated marginal mean changes from baseline to 6 months (95% CI) from linear mixed-effects models with a random intercept per participant, adjusted for age, sex, body mass index and baseline antihypertensive medication status (each with a time interaction), time of day and season. Sex-stratified models omit sex terms. Full cohort additionally includes participants with normal baseline BP. n, participants contributing to the adjusted SBP model. No recorded antihypertensive medication: no journal entry from 180 days before baseline through the 6-month reading. No recorded GLP-1 receptor agonist use: no journal entry from 30 days before baseline through the 6-month reading. Excluding antihypertensive initiation during follow-up: excludes participants whose first logged antihypertensive medication was after baseline. BP, blood pressure; CI, confidence interval; DBP, diastolic blood pressure; GLP-1, glucagon-like peptide-1; SBP, systolic blood pressure.

**Table S4. Blood pressure change from baseline at 3, 6 and 12 months by baseline blood pressure category, from the model including the 12-month timepoint**

| Stratum | Time (Months) | n | Unadjusted Δ (mmHg) | Adjusted Δ (mmHg) | RTM-corrected Δ (mmHg) | P | d |
|---|---|---|---|---|---|---|---|
| **Systolic Blood Pressure** | | | | | | | |
| Elevated | 3 | 2,564 | −3.0 (−3.3, −2.6) | −2.8 (−3.3, −2.3) | — | <0.001 | 0.28 |
| | 6 | 5,163 | −2.5 (−2.8, −2.2) | −2.6 (−3.1, −2.2) | — | <0.001 | 0.26 |
| | 12 | 421 | −3.8 (−4.7, −3.0) | −4.4 (−5.7, −3.1) | −4.6 (−5.9, −3.2) | <0.001 | 0.44 |
| Stage 1 | 3 | 5,279 | −4.1 (−4.4, −3.9) | −4.1 (−4.6, −3.7) | — | <0.001 | 0.41 |
| | 6 | 10,269 | −3.7 (−3.9, −3.5) | −4.0 (−4.3, −3.6) | — | <0.001 | 0.39 |
| | 12 | 712 | −5.3 (−5.9, −4.7) | −5.9 (−7.1, −4.8) | −4.8 (−6.0, −3.4) | <0.001 | 0.59 |
| Stage 2 | 3 | 2,403 | −10.6 (−11.0, −10.2) | −10.8 (−11.4, −10.3) | — | <0.001 | 1.08 |
| | 6 | 4,571 | −11.6 (−11.9, −11.3) | −12.1 (−12.5, −11.7) | — | <0.001 | 1.21 |
| | 12 | 284 | −14.4 (−15.4, −13.4) | −15.3 (−16.7, −13.9) | −9.2 (−10.8, −7.5) | <0.001 | 1.53 |
| Marginal $R^2$ | | | 0.25 | 0.28 | | | |
| Conditional $R^2$ | | | 0.52 | 0.53 | | | |
| Residual Skew/Excess Kurtosis | | | 0.24 / 2.2 | 0.23 / 2.3 | | | |
| **Diastolic Blood Pressure** | | | | | | | |
| Elevated | 3 | 2,564 | 0.3 (0.0, 0.6) | 0.1 (−0.3, 0.5) | — | 0.654 | 0.01 |
| | 6 | 5,163 | 0.7 (0.5, 0.9) | 0.3 (−0.1, 0.6) | — | 0.322 | 0.03 |
| | 12 | 421 | −0.2 (−0.8, 0.4) | −0.7 (−1.7, 0.3) | −2.3 (−3.5, −1.1) | 0.322 | 0.08 |
| Stage 1 | 3 | 5,279 | −3.2 (−3.4, −3.0) | −3.5 (−3.9, −3.2) | — | <0.001 | 0.42 |
| | 6 | 10,269 | −3.2 (−3.4, −3.1) | −3.7 (−4.0, −3.5) | — | <0.001 | 0.44 |
| | 12 | 712 | −4.0 (−4.4, −3.5) | −4.5 (−5.4, −3.7) | −3.4 (−4.5, −2.2) | <0.001 | 0.54 |
| Stage 2 | 3 | 2,403 | −7.5 (−7.8, −7.2) | −8.0 (−8.4, −7.6) | — | <0.001 | 0.96 |
| | 6 | 4,571 | −8.4 (−8.7, −8.2) | −9.2 (−9.5, −8.8) | — | <0.001 | 1.09 |
| | 12 | 284 | −10.3 (−11.0, −9.6) | −11.1 (−12.2, −10.1) | −7.1 (−8.4, −5.7) | <0.001 | 1.33 |
| Marginal $R^2$ | | | 0.33 | 0.34 | | | |
| Conditional $R^2$ | | | 0.58 | 0.58 | | | |
| Residual Skew/Excess Kurtosis | | | 0.22 / 1.6 | 0.23 / 1.6 | | | |

Values are estimated marginal mean changes (mmHg) from baseline with 95% confidence intervals from linear mixed-effects models with a participant random intercept fitted to baseline, 3-, 6- and 12-month readings; the adjusted model includes age, sex, baseline BMI and baseline antihypertensive medication status, each interacted with time, plus time of day and season. RTM-corrected Δ adds the expected regression to the mean to the adjusted Δ, with 95% percentile confidence intervals from 2,000 participant-level bootstrap resamples; at 12 months the reference mean and variance are taken from participants with a 12-month reading. n, participants with a baseline and a same-timepoint reading. P values are Holm-corrected within the within-stratum contrast family and, like d, come from this four-timepoint model, so 3- and 6-month values differ slightly from Table 2. d, absolute adjusted Δ divided by the pooled baseline SD. BMI, body mass index; RTM, regression to the mean. $R^2$, Nakagawa marginal and conditional $R^2$ of the unadjusted and adjusted models; residual skew and excess kurtosis are from the same models.

**Table S5. Associations of platform engagement with a ≥5 mmHg reduction in blood pressure at 6 months in the primary cohort and in sensitivity cohorts**

| Engagement metric | Primary cohort | No recorded antihypertensive medication | No recorded GLP-1 receptor agonist use | Excluding antihypertensive initiation during follow-up |
|---|---|---|---|---|
| **≥5 mmHg reduction in SBP** | | | | |
| Participants, n | 19,950–19,986 | 16,274–16,305 | 19,240–19,275 | 18,424–18,458 |
| Daily app time | 1.30 (1.25, 1.36)* | 1.32 (1.26, 1.39)* | 1.29 (1.24, 1.35)* | 1.30 (1.24, 1.36)* |
| Journal completion | 1.02 (1.02, 1.03)* | 1.02 (1.01, 1.03)* | 1.02 (1.01, 1.03)* | 1.02 (1.01, 1.03)* |
| Notification engagement | 1.04 (1.03, 1.05)* | 1.05 (1.03, 1.06)* | 1.04 (1.03, 1.05)* | 1.04 (1.03, 1.05)* |
| AI Coach engagement | 1.07 (1.06, 1.09)* | 1.08 (1.06, 1.10)* | 1.07 (1.05, 1.09)* | 1.07 (1.06, 1.09)* |
| Sleep need achieved | 1.09 (1.05, 1.14)* | 1.09 (1.04, 1.14)* | 1.10 (1.05, 1.14)* | 1.09 (1.05, 1.14)* |
| Physical activity recommendation | 1.01 (0.98, 1.03) | 1.01 (0.99, 1.04) | 1.01 (0.98, 1.03) | 1.00 (0.98, 1.03) |
| Blood pressure readings | 1.28 (1.22, 1.33)* | 1.30 (1.24, 1.37)* | 1.27 (1.22, 1.33)* | 1.30 (1.24, 1.36)* |
| **≥5 mmHg reduction in DBP** | | | | |
| Participants, n | 19,950–19,986 | 16,274–16,305 | 19,240–19,275 | 18,424–18,458 |
| Daily app time | 1.35 (1.29, 1.41)* | 1.36 (1.29, 1.43)* | 1.33 (1.27, 1.39)* | 1.34 (1.28, 1.40)* |
| Journal completion | 1.02 (1.01, 1.03)* | 1.02 (1.01, 1.03)* | 1.02 (1.01, 1.03)* | 1.02 (1.01, 1.03)* |
| Notification engagement | 1.04 (1.03, 1.05)* | 1.04 (1.03, 1.05)* | 1.04 (1.03, 1.05)* | 1.04 (1.03, 1.05)* |
| AI Coach engagement | 1.08 (1.06, 1.09)* | 1.08 (1.06, 1.10)* | 1.07 (1.06, 1.09)* | 1.07 (1.06, 1.09)* |
| Sleep need achieved | 1.16 (1.11, 1.21)* | 1.18 (1.12, 1.23)* | 1.16 (1.12, 1.21)* | 1.17 (1.12, 1.22)* |
| Physical activity recommendation | 1.02 (1.00, 1.05) | 1.02 (0.99, 1.05) | 1.03 (1.00, 1.05) | 1.02 (0.99, 1.05) |
| Blood pressure readings | 1.14 (1.09, 1.19)* | 1.14 (1.08, 1.20)* | 1.13 (1.08, 1.19)* | 1.15 (1.09, 1.20)* |

Values are odds ratios (95% profile-likelihood CI) from separate logistic regression models, one engagement metric per model, adjusted for baseline BP of the same outcome, age, sex, body mass index and baseline antihypertensive medication status (omitted in the cohort with no recorded antihypertensive medication, where it is constant). Odds ratios are per 10 percentage points of days with wearable data for journal completion, notification engagement and AI Coach engagement; per 10 percentage points of days with a Strain recommendation for the physical activity recommendation; per 10 percentage points of nightly sleep need achieved; and per 1-unit increase in log(app minutes per day of app use + 1) and log(number of BP readings + 1). *$P < 0.005$, the prespecified significance threshold. n ranges across metrics because each model uses complete cases for its metric. No recorded antihypertensive medication: no journal entry from 180 days before baseline through the 6-month reading. No recorded GLP-1 receptor agonist use: no journal entry from 30 days before baseline through the 6-month reading. Excluding antihypertensive initiation during follow-up: excludes participants whose first logged antihypertensive medication was after baseline. BP, blood pressure; CI, confidence interval; DBP, diastolic blood pressure; GLP-1, glucagon-like peptide-1; SBP, systolic blood pressure.

**Table S6. Associations of baseline and 6-month wearable-derived metrics with a ≥5 mmHg reduction in blood pressure at 6 months in the primary cohort and in sensitivity cohorts**

| | Primary cohort | Full cohort (including normal BP) | No recorded antihypertensive medication | No recorded GLP-1 receptor agonist use | Excluding antihypertensive initiation during follow-up |
|---|---|---|---|---|---|
| **≥5 mmHg reduction in systolic blood pressure** | | | | | |
| Participants (responders), n | 14,189 (7,051) | 20,360 (8,214) | 11,831 (5,914) | 13,698 (6,779) | 13,751 (6,808) |
| RHR, baseline | 1.026 (1.010, 1.042)* | 1.027 (1.012, 1.041)* | 1.022 (1.004, 1.040) | 1.028 (1.012, 1.045)* | 1.025 (1.009, 1.042)* |
| RHR, 6-month | 0.973 (0.958, 0.989)* | 0.974 (0.960, 0.988)* | 0.976 (0.958, 0.994) | 0.970 (0.954, 0.986)* | 0.975 (0.959, 0.991)* |
| HRV, baseline | 1.000 (0.994, 1.006) | 1.001 (0.995, 1.006) | 0.999 (0.993, 1.006) | 1.000 (0.994, 1.006) | 0.999 (0.993, 1.005) |
| HRV, 6-month | 1.000 (0.994, 1.006) | 0.999 (0.994, 1.005) | 1.001 (0.994, 1.008) | 1.000 (0.994, 1.006) | 1.001 (0.995, 1.007) |
| Sleep duration, baseline | 1.008 (0.908, 1.120) | 1.009 (0.919, 1.108) | 0.970 (0.863, 1.092) | 1.015 (0.912, 1.130) | 1.016 (0.913, 1.132) |
| Sleep duration, 6-month | 1.033 (0.921, 1.160) | 1.040 (0.938, 1.153) | 1.088 (0.957, 1.238) | 1.024 (0.910, 1.151) | 1.031 (0.917, 1.160) |
| Sleep consistency, baseline | 0.996 (0.990, 1.003) | 0.994 (0.988, 0.999) | 0.998 (0.991, 1.005) | 0.997 (0.991, 1.004) | 0.997 (0.991, 1.003) |
| Sleep consistency, 6-month | 1.006 (0.999, 1.014) | 1.010 (1.004, 1.017)* | 1.004 (0.996, 1.012) | 1.006 (0.999, 1.013) | 1.006 (0.999, 1.013) |
| Physical activity, baseline | 0.986 (0.963, 1.010) | 0.986 (0.966, 1.007) | 0.976 (0.952, 1.002) | 0.989 (0.966, 1.013) | 0.984 (0.961, 1.008) |
| Physical activity, 6-month | 1.019 (0.990, 1.048) | 1.015 (0.990, 1.040) | 1.030 (0.999, 1.063) | 1.014 (0.985, 1.044) | 1.023 (0.994, 1.053) |
| **≥5 mmHg reduction in diastolic blood pressure** | | | | | |
| Participants (responders), n | 14,189 (5,702) | 20,360 (6,809) | 11,831 (4,781) | 13,698 (5,466) | 13,751 (5,504) |
| RHR, baseline | 1.023 (1.007, 1.040)* | 1.023 (1.008, 1.037)* | 1.017 (0.999, 1.036) | 1.025 (1.008, 1.042)* | 1.023 (1.006, 1.040) |
| RHR, 6-month | 0.966 (0.950, 0.982)* | 0.967 (0.952, 0.981)* | 0.971 (0.953, 0.989)* | 0.964 (0.947, 0.981)* | 0.967 (0.951, 0.984)* |
| HRV, baseline | 0.997 (0.991, 1.004) | 1.000 (0.994, 1.005) | 0.995 (0.988, 1.001) | 0.998 (0.991, 1.004) | 0.997 (0.991, 1.003) |
| HRV, 6-month | 1.004 (0.998, 1.011) | 1.002 (0.997, 1.008) | 1.007 (1.000, 1.014) | 1.004 (0.998, 1.011) | 1.005 (0.998, 1.011) |
| Sleep duration, baseline | 0.916 (0.822, 1.022) | 0.948 (0.860, 1.045) | 0.865 (0.766, 0.978) | 0.920 (0.823, 1.028) | 0.914 (0.817, 1.022) |
| Sleep duration, 6-month | 1.232 (1.093, 1.389)* | 1.194 (1.073, 1.329)* | 1.304 (1.141, 1.491)* | 1.229 (1.087, 1.389)* | 1.243 (1.100, 1.406)* |
| Sleep consistency, baseline | 0.999 (0.992, 1.005) | 0.996 (0.990, 1.001) | 1.000 (0.993, 1.007) | 0.998 (0.992, 1.005) | 0.999 (0.993, 1.006) |
| Sleep consistency, 6-month | 1.004 (0.996, 1.011) | 1.006 (1.000, 1.013) | 1.004 (0.996, 1.012) | 1.004 (0.996, 1.012) | 1.003 (0.995, 1.011) |
| Physical activity, baseline | 0.950 (0.927, 0.974)* | 0.943 (0.923, 0.963)* | 0.948 (0.923, 0.974)* | 0.953 (0.929, 0.977)* | 0.951 (0.928, 0.975)* |
| Physical activity, 6-month | 1.081 (1.050, 1.114)* | 1.090 (1.062, 1.118)* | 1.088 (1.053, 1.124)* | 1.075 (1.043, 1.108)* | 1.081 (1.049, 1.114)* |

Values are odds ratios (95% profile-likelihood CI) per unit increase in the metric, from one multivariable logistic regression per outcome and cohort containing all ten metrics (30-day pre-baseline mean and 6-month study-window mean of RHR, HRV, sleep duration, sleep consistency and physical activity), adjusted for baseline BP of the same outcome, age, sex, body mass index and baseline antihypertensive medication status (omitted in the cohort with no recorded antihypertensive medication, where it is constant). Units: RHR, beats per minute; HRV, ms; sleep duration, hours; sleep consistency, points on a 0–100 scale; physical activity, Strain units on a 0–21 scale. Models require wearable data on at least 8 of 30 baseline days. Participants (responders), participants in the model and the number with a ≥5 mmHg reduction. *P < 0.005, the prespecified significance threshold. Full cohort additionally includes participants with normal baseline BP. No recorded antihypertensive medication: no journal entry from 180 days before baseline through the 6-month reading. No recorded GLP-1 receptor agonist use: no journal entry from 30 days before baseline through the 6-month reading. Excluding antihypertensive initiation during follow-up: excludes participants whose first logged antihypertensive medication was after baseline. BP, blood pressure; CI, confidence interval; GLP-1, glucagon-like peptide-1; HRV, heart rate variability; RHR, resting heart rate.

**Table S7. Between- and within-person associations of wearable-derived metrics with blood pressure at the reading level in the primary cohort and in sensitivity analyses**

| | Primary cohort | Raw BP outcome | Full cohort (including normal BP) | No recorded antihypertensive medication | No recorded GLP-1 receptor agonist use | Excluding antihypertensive initiation during follow-up |
|---|---|---|---|---|---|---|
| **Systolic blood pressure** | | | | | | |
| Participants (readings), n | 16,161 (132,843) | 16,161 (132,843) | 22,408 (185,310) | 12,893 (97,586) | 15,543 (126,955) | 14,765 (116,813) |
| RHR, between-person | −0.03 (−0.22, 0.16) | −0.03 (−0.18, 0.13) | −0.12 (−0.29, 0.05) | −0.03 (−0.24, 0.19) | 0.06 (−0.14, 0.25) | −0.04 (−0.24, 0.16) |
| HRV, between-person | 0.10 (−0.07, 0.27) | −0.01 (−0.15, 0.13) | 0.17 (0.02, 0.32) | 0.08 (−0.11, 0.27) | 0.11 (−0.06, 0.28) | 0.06 (−0.11, 0.24) |
| Sleep duration, between-person | −0.18 (−0.34, −0.02) | −0.22 (−0.35, −0.09)* | −0.20 (−0.34, −0.06)* | −0.19 (−0.36, −0.01) | −0.17 (−0.34, −0.01) | −0.17 (−0.34, −0.01) |
| Sleep consistency, between-person | −0.10 (−0.26, 0.05) | −0.26 (−0.39, −0.13)* | −0.12 (−0.26, 0.01) | −0.12 (−0.29, 0.05) | −0.10 (−0.26, 0.06) | −0.12 (−0.29, 0.04) |
| Physical activity, between-person | −0.37 (−0.53, −0.21)* | −0.33 (−0.46, −0.19)* | −0.30 (−0.44, −0.15)* | −0.39 (−0.57, −0.21)* | −0.37 (−0.53, −0.20)* | −0.41 (−0.57, −0.24)* |
| RHR, within-person | 0.56 (0.47, 0.65)* | 0.56 (0.47, 0.65)* | 0.50 (0.43, 0.57)* | 0.59 (0.49, 0.70)* | 0.56 (0.47, 0.65)* | 0.56 (0.47, 0.66)* |
| HRV, within-person | 0.03 (−0.05, 0.11) | 0.03 (−0.05, 0.11) | 0.01 (−0.05, 0.07) | 0.06 (−0.02, 0.14) | 0.03 (−0.05, 0.10) | 0.06 (−0.02, 0.14) |
| Sleep duration, within-person | −0.13 (−0.19, −0.08)* | −0.13 (−0.19, −0.08)* | −0.11 (−0.16, −0.07)* | −0.13 (−0.20, −0.06)* | −0.11 (−0.17, −0.05)* | −0.13 (−0.19, −0.07)* |
| Sleep consistency, within-person | −0.03 (−0.09, 0.02) | −0.03 (−0.09, 0.02) | −0.03 (−0.08, 0.01) | −0.04 (−0.09, 0.02) | −0.03 (−0.08, 0.02) | −0.03 (−0.08, 0.03) |
| Physical activity, within-person | −0.18 (−0.24, −0.12)* | −0.18 (−0.24, −0.12)* | −0.16 (−0.21, −0.11)* | −0.18 (−0.25, −0.12)* | −0.19 (−0.25, −0.13)* | −0.19 (−0.25, −0.13)* |
| **Diastolic blood pressure** | | | | | | |
| Participants (readings), n | 16,161 (132,843) | 16,161 (132,843) | 22,408 (185,310) | 12,893 (97,586) | 15,543 (126,955) | 14,765 (116,813) |
| RHR, between-person | −0.29 (−0.43, −0.15)* | 0.61 (0.49, 0.73)* | −0.24 (−0.36, −0.11)* | −0.32 (−0.48, −0.16)* | −0.23 (−0.37, −0.09)* | −0.29 (−0.43, −0.14)* |
| HRV, between-person | 0.02 (−0.11, 0.15) | −0.15 (−0.26, −0.04) | 0.02 (−0.09, 0.13) | −0.01 (−0.16, 0.14) | 0.01 (−0.12, 0.14) | 0.01 (−0.12, 0.14) |
| Sleep duration, between-person | −0.10 (−0.22, 0.02) | −0.21 (−0.30, −0.11)* | −0.12 (−0.22, −0.02) | −0.11 (−0.25, 0.02) | −0.10 (−0.22, 0.02) | −0.11 (−0.23, 0.02) |
| Sleep consistency, between-person | 0.00 (−0.12, 0.13) | −0.11 (−0.21, −0.02) | −0.01 (−0.12, 0.09) | 0.03 (−0.11, 0.17) | −0.01 (−0.13, 0.12) | 0.01 (−0.12, 0.14) |
| Physical activity, between-person | −0.41 (−0.54, −0.28)* | −0.47 (−0.57, −0.37)* | −0.33 (−0.44, −0.22)* | −0.43 (−0.57, −0.29)* | −0.40 (−0.53, −0.27)* | −0.43 (−0.56, −0.30)* |
| RHR, within-person | 0.59 (0.53, 0.66)* | 0.59 (0.53, 0.66)* | 0.53 (0.48, 0.58)* | 0.59 (0.51, 0.66)* | 0.58 (0.52, 0.65)* | 0.58 (0.52, 0.65)* |
| HRV, within-person | 0.01 (−0.04, 0.07) | 0.01 (−0.04, 0.07) | 0.00 (−0.04, 0.04) | 0.04 (−0.02, 0.09) | 0.01 (−0.04, 0.06) | 0.03 (−0.02, 0.08) |
| Sleep duration, within-person | −0.07 (−0.11, −0.03)* | −0.07 (−0.11, −0.03)* | −0.06 (−0.09, −0.03)* | −0.06 (−0.10, −0.01) | −0.05 (−0.09, −0.01) | −0.06 (−0.10, −0.02) |
| Sleep consistency, within-person | −0.03 (−0.06, 0.01) | −0.03 (−0.06, 0.01) | −0.04 (−0.07, −0.01) | −0.04 (−0.08, 0.00) | −0.02 (−0.06, 0.01) | −0.03 (−0.07, 0.01) |
| Physical activity, within-person | −0.28 (−0.32, −0.23)* | −0.28 (−0.32, −0.23)* | −0.26 (−0.30, −0.22)* | −0.29 (−0.34, −0.25)* | −0.28 (−0.33, −0.24)* | −0.30 (−0.34, −0.26)* |

Values are β (95% CI) in mmHg per 1 SD of the predictor from linear mixed-effects models of each non-baseline BP reading's deviation from the participant's baseline BP (raw BP outcome: the reading itself, with baseline BP as a covariate), with a participant random intercept and cluster-robust (CR1) standard errors, adjusted for age, sex, body mass index and baseline antihypertensive medication status (omitted in the cohort with no recorded antihypertensive medication, where it is constant). Each metric is the 7-day look-back mean, decomposed into a between-person component (participant mean across non-baseline readings) and a within-person component (reading-level deviation from that mean), each standardised to its SD. Participants contribute at least two non-baseline readings. Participants (readings), participants and BP readings in the model. *P < 0.005, the prespecified significance threshold. Full cohort additionally includes participants with normal baseline BP. No recorded antihypertensive medication: no journal entry from 180 days before baseline through the 6-month reading. No recorded GLP-1 receptor agonist use: no journal entry from 30 days before baseline through the 6-month reading. Excluding antihypertensive initiation during follow-up: excludes participants whose first logged antihypertensive medication was after baseline. BP, blood pressure; CI, confidence interval; GLP-1, glucagon-like peptide-1; HRV, heart rate variability; RHR, resting heart rate; SD, standard deviation.
